# Risk of Cardiovascular Events Using the SMART Polyvascular Disease Risk Score

**DOI:** 10.1101/2025.09.05.25335208

**Authors:** Subhash Banerjee, Anand Gupta, Minseob Jeong, David Fernandez Vazquez, Shuaib Abdullah, Bradley R. Grimsley, Rohit J. Parmar, Vishal Ahuja, Robert C. Stoler, Ambarish Pandey

## Abstract

**Background:** Secondary Manifestations of Arterial Disease (SMART) risk score based projection of risk for cardiovascular (CV) events can help devise risk mitigation strategies.

**Objective:** Use of the SMART polyvascular disease score to compute the risk for major adverse cardiovascular events (MACE) in U.S. patients.

**Methods:** We accessed the Baylor Scott & White EPIC informatics and data warehouse to identify patients at their first outpatient cardiology evaluation between April 2014 and October 2023 to estimate up to 10-year risk of MACE, a composite of all-cause death, ischemic stroke, and non-fatal myocardial infarction (MI). Cox regression, accelerated failure time model, and survival analyses were used to develop and validate the SMART risk score.

**Results:** The study population of 259,250 patients (mean age 60.9 ± 15.2 years, 48.6% female) were divided into development (60%) and test (40%) cohorts; median follow-up 2.1 years (interquartile range 0.54 - 4.4). The SMART risk score allowed accurate estimation of MACE. Patients in low (<10%), moderate (10% - <20%), high (20% - <30%), and very high (≥30%) SMART risk score groups had observed MACE events rates of 2.9%, 15.0%, 24.5% and 56.5%, respectively in the test cohort (p<0.0001 for all inter-group comparisons). Most MACE events were all-cause death, with nonfatal MI and stroke also being high, in the very high-risk group. The SMART score outperformed an established risk prediction model (c-statistic=0.811) in the test cohort.

**Conclusion:** The SMART Polyvascular disease risk score can provide accurate estimation of up to 10-year risk of CV events and could be potentially leveraged to develop individualized risk mitigation strategies.

## Introduction

The Secondary Manifestations of Arterial Disease (SMART) risk score, developed and validated by Dorresteijn JA et al., was designed to estimate the risk of recurrent cardiovascular events in patients with pre-existing cardiovascular disease (CVD).^1^ The SMART risk prediction model is an established calculator to estimate the 10-year risk of non-fatal myocardial infarction (MI), ischemic stroke, or all-cause death, collectively termed as major adverse cardiovascular events or MACE from readily available clinical characteristics.^1^

Recently, our group published an application and validation of the SMART risk score in a cohort of U.S. Veterans.^2^ The study aimed to redefine cardiovascular risk assessment by shifting the focus from a binary category of secondary prevention to a continuous risk spectrum, utilizing the SMART risk score.^3^ The overarching goal was to evaluate a structured risk assessment strategy to guide potential interventions for lowering the risk of recurrent cardiovascular events. To this end, the SMART risk score, which integrates 14 easily obtainable clinical and laboratory parameters, proved to be a highly suitable tool.^1^ The variables used in computing the SMART risk score include age, sex, smoking, time (in years) since CVD diagnoses to cardiology visit and clinical parameters [(average systolic blood pressure, history of diabetes mellitus (DM), coronary artery disease (CAD), cerebrovascular disease, abdominal aortic aneurysm (AAA), peripheral artery disease (PAD), high-density lipoprotein (HDL) cholesterol, total cholesterol (TC), estimated glomerular filtration rate (eGFR), and high-sensitivity C-reactive protein (hsCRP)].^1, 2^

The aim of the current study was to apply the SMART risk score to an all-comer outpatient U.S. non-Veteran patient population in order to predict 10-year MACE, a composite of all-cause death, MI, and ischemic stroke.

## Methods

### Study Population

The study included patients from the Baylor Scott & White Health (BSWH) system, which comprises 52 hospitals and over 1,300 sites of care. Between April 1, 2014, and October 31, 2023, a total of 3,461,702 patients visited the health system, as recorded in the EPIC electronic health records (EHR). We analyzed 302,689 adult patients (≥18 years) who were referred to cardiology for atherosclerotic cardiovascular disease (ASCVD), including cerebrovascular disease, CAD, PAD, AAA, or polyvascular disease states. Patients without a recorded time since first CVD diagnosis were excluded. Additionally, those with extreme outliers in continuous variables were removed. **Figure 1** illustrates formation of the study cohort. The time to first MACE was assessed for patients who experienced a MACE event following their initial cardiology visit. Patients who did not experience a MACE event and lacked 10-year follow-up data were considered left-censored. Patients who remained free of MACE for up to 10 years after their cardiology visit were considered right censored. The final study cohort included 259,250 patients. The dataset was split into a development cohort (60%, n = 155,550) and a test cohort (40%, n = 103,700). This study was approved by the Baylor University Medical Center institutional review board.

**Figure 1:**
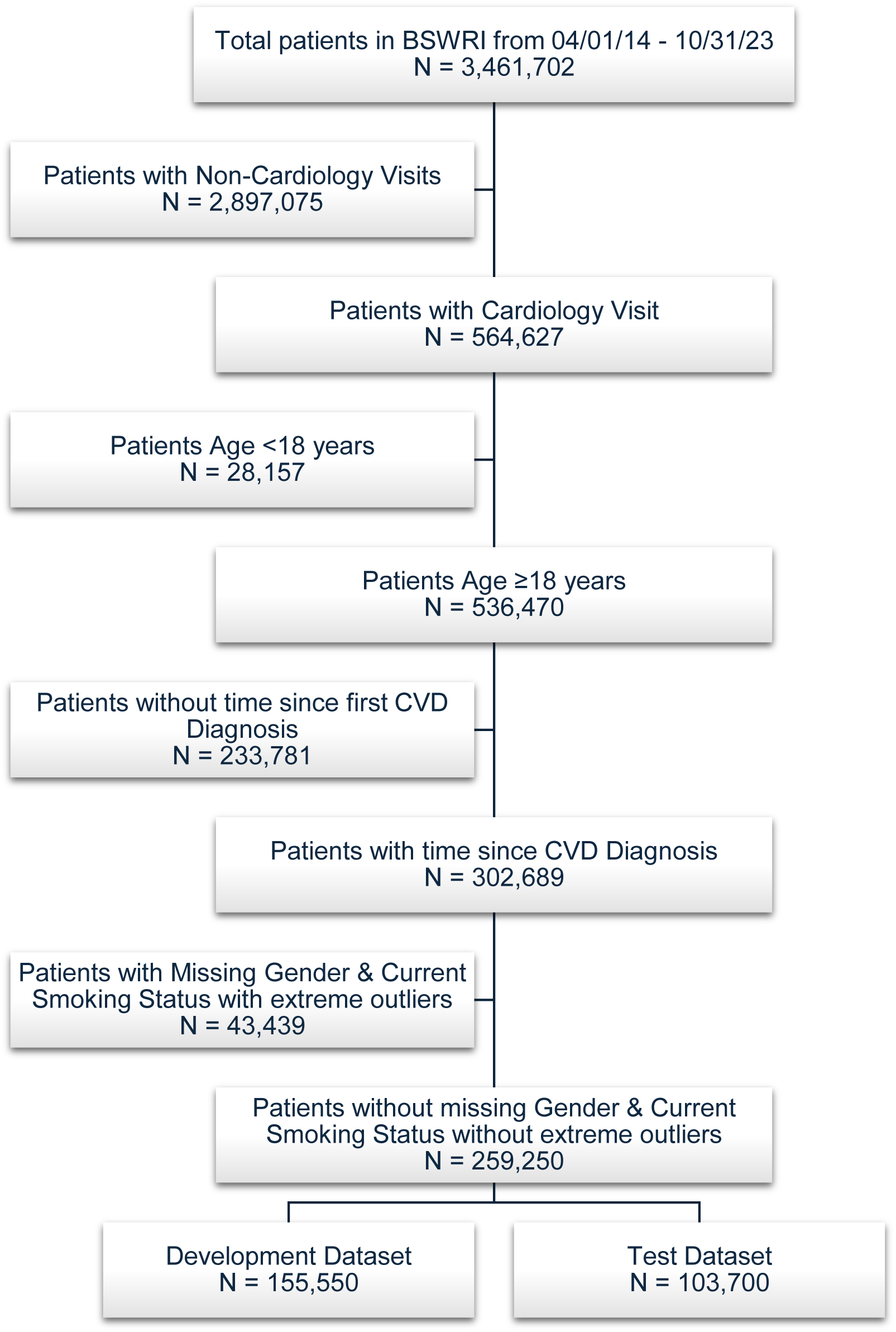
Study Cohort: Describes development of the study cohort. ICD – international classification of disease (includes ICD 9 and 10); CPT – current procedural terminology.

### Key covariates of interest for SMART Risk score

The present study was conducted to evaluate the performance and applicability of the SMART risk score in the BSWH cohort. The SMART risk score variables included in the model were age, sex, current smoking status, time in years since CVD diagnosis to the first cardiology visit, systolic blood pressure, history of DM, CAD, cerebrovascular disease, AAA, PAD, HDL, TC, eGFR, and hsCRP.^1, 2^ Patient age was calculated as the difference between the initial cardiology visit and the date of birth. Sex, Current smoking status and systolic blood pressure was extracted at the date of the cardiology visit. Comorbidities like DM, CAD, cerebrovascular disease, AAA, PAD and CVD were defined using International Classification of Diseases (ICD) 9 and 10 and Current Procedure Terminology (CPT) codes **(Appendix 1)**. TRIPOD guideline adherence document is included as **Appendix 2.** Patient’s time since CVD diagnosis is the first time the patient was diagnosed with CVD before the initial cardiology visits and was obtained from clinical records. Laboratory data were obtained within 365 days of the initial cardiology. All data were directly extracted via EPIC Clarity.

### Outcome of interest

The primary outcome was MACE, a composite of all-cause death, ischemic stroke, and non-fatal MI, within 10 years of the first cardiology visit. MI or ischemic stroke occurring after the initial cardiology visit and before the recorded date of death (if any) were defined using ICD 9 and 10 and CPT codes. All-cause death was identified from EPIC records, including in-hospital deaths and outpatient updates documented after follow-up calls or when clinics were informed. Outcomes were assessed over a 10-year period from the date of the first cardiology visit. A MACE event was considered for the earliest of the three outcomes. Censoring was applied at the earliest of the following events: the date of last documented follow-up or the completion of the 10-year follow-up window.

### Statistical Analysis

Descriptive analysis was performed on the overall cohort. Missing values were imputed separately for each dataset after splitting. Initially, approximately 0.57% of patients had hsCRP values, and 3.22% had C-reactive protein (CRP) values. Imputing hsCRP values was a multistep process. First, a linear regression model was used to estimate hsCRP from CRP values giving us a total of 3.79% hsCRP values. Second, the total 3.79% hsCRP values, systolic blood pressure, eGFR, TC and HDL were imputed by identifying the extent of missing data assessed using Little’s Missing Completely at Random (MCAR) test (statistic=29,929.66, p<0.05), which suggested the data were likely missing at random (MAR).^2, 4^ Multiple Imputation by Chained Equations (MICE) was used, enhanced by predictive mean matching, ensuring plausible and consistent imputations for the variables. ^5, 6^

Levene’s test was performed to assess the homogeneity of error variances. Continuous predictors were trimmed at the 1st and 99th percentiles to limit the impact of outliers. The Cox proportional hazards model was initially explored; however, repeated assessments revealed violations of the proportional hazard’s assumption across multiple covariates. Although, in the Accelerated Failure Time (AFT) framework, the reported hazard ratios (HRs) reflect the extent to which covariates accelerate or decelerate the time to MACE. ^7, 8^ As a result, an AFT model assuming a Weibull distribution was employed to model time to MACE, providing a more appropriate framework for estimating the impact of covariates on event timing. The development dataset was used to build the SMART risk model, which was then validated in the test cohort.

### SMART Risk score model evaluation

Considering the differences in the primary cohort where SMART risk score was derived and the intended use population from BSW health system, we did not use the out-of-the box estimates for the model covariates. Instead, model optimization was performed in the randomly selected 60% study development cohort. For this, natural log transformations were applied to the variables “time since CVD diagnosis to cardiology visit” and “hsCRP,” as they initially did not meet the linearity assumption. Maximum likelihood estimation was used to fit the AFT model. Patients who remained event-free for 10 years were right-censored, while those with less than 10 years of follow-up were left-censored. Model fit was evaluated using deviance residuals to identify potential outliers and assess overall model adequacy. Model performance was further assessed by examining the log-likelihood, likelihood ratio chi-square statistics, and Akaike Information Criterion (AIC) across candidate models to guide model selection and identify the best-fitting approach.

The performance of the SMART risk score was evaluated in the 40% randomly selected test cohort. Discrimination was assessed using the concordance (C)-statistics, calculated separately for the development, test, and overall cohorts. Calibration was examined through calibration plots comparing predicted versus observed risks. The D’Agostino Nam test used to test the deviance of the predicted from the observed outcomes. Clinical significance of the deviation was accessed based on the calibration plot **(Figure 2.)**. Kaplan–Meier (K-M) survival estimates were calculated for the risk of MACE across the SMART risk score strata and compared using Log-rank, Breslow, and Tarone-Ware tests. Clinical utility was assessed using decision curve analysis, which quantified the net benefit of prediction-guided decision-making across a range of threshold probabilities. Comparative performance between the previously established TRS2°P score and the SMART risk score was examined using decision curve analysis, Brier scores, area under the curve (AUC), and concordance statistics.^3^ Additionally, an alternative model excluding hsCRP was constructed to evaluate model performance in the absence of this biomarker. Subgroup analyses stratified by sex, race, and ethnicity were performed across the full cohort to assess the robustness and generalizability of the findings. All analyses were performed using SQL and R v.4.2.2 (http://www.R-project.org) with relevant statistical packages.

**Figure 2:**
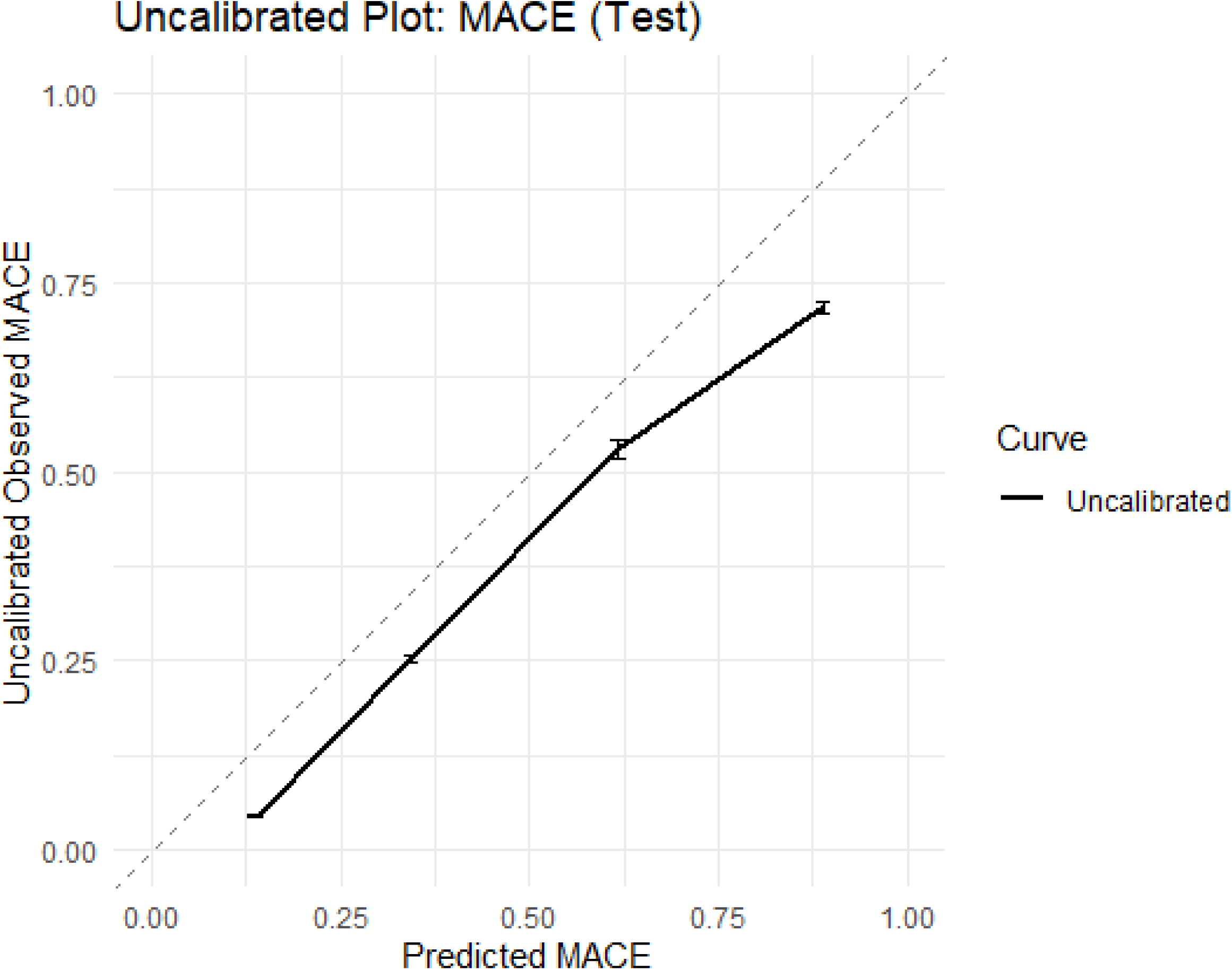
Uncalibrated Plot for the Test Cohort: MACE – major adverse cardiovascular event.

## RESULTS

Baseline characteristics of 259,250 patients included in this analysis were stratified into SMART risk score low, moderate, high, and very-high risk groups are shown as **Table 1**. Nearly 48.6% patients included were women. The mean age was 60.9 ± 15.2, with higher mean age in high (69.9 ± 9.7) and very high risk (72.2 ± 9.97) groups. Majority of the patients were white (82.2%), and African Americans constituting nearly 10.9%. The prevalence of DM, CAD, cerebrovascular disease, PAD, and AAA were expectedly higher in the high and very high-risk groups. Mean values for TC, LDL, and eGFR were 173.4 ± 47.1 mg/dL, 102.5 ± 36.5 mg/dL, and 81.5 ± 25.5 mL/min/1.73 m^2^, respectively. The entire study cohort (n=259,250) was randomly divided into development (n=155,550) and test cohorts (n=103,700) using random sampling without replacement. All baseline variables were statistically significantly different between each of the groups (low, moderate & high risk) versus very high-risk group. **Supplementary Tables 1 and 2** illustrate the baseline characteristics of the development and test cohorts divided into the four SMART risk groups. The median follow-up time was 2.1 years (interquartile range 0.54–4.4 years).

**Table 1:**
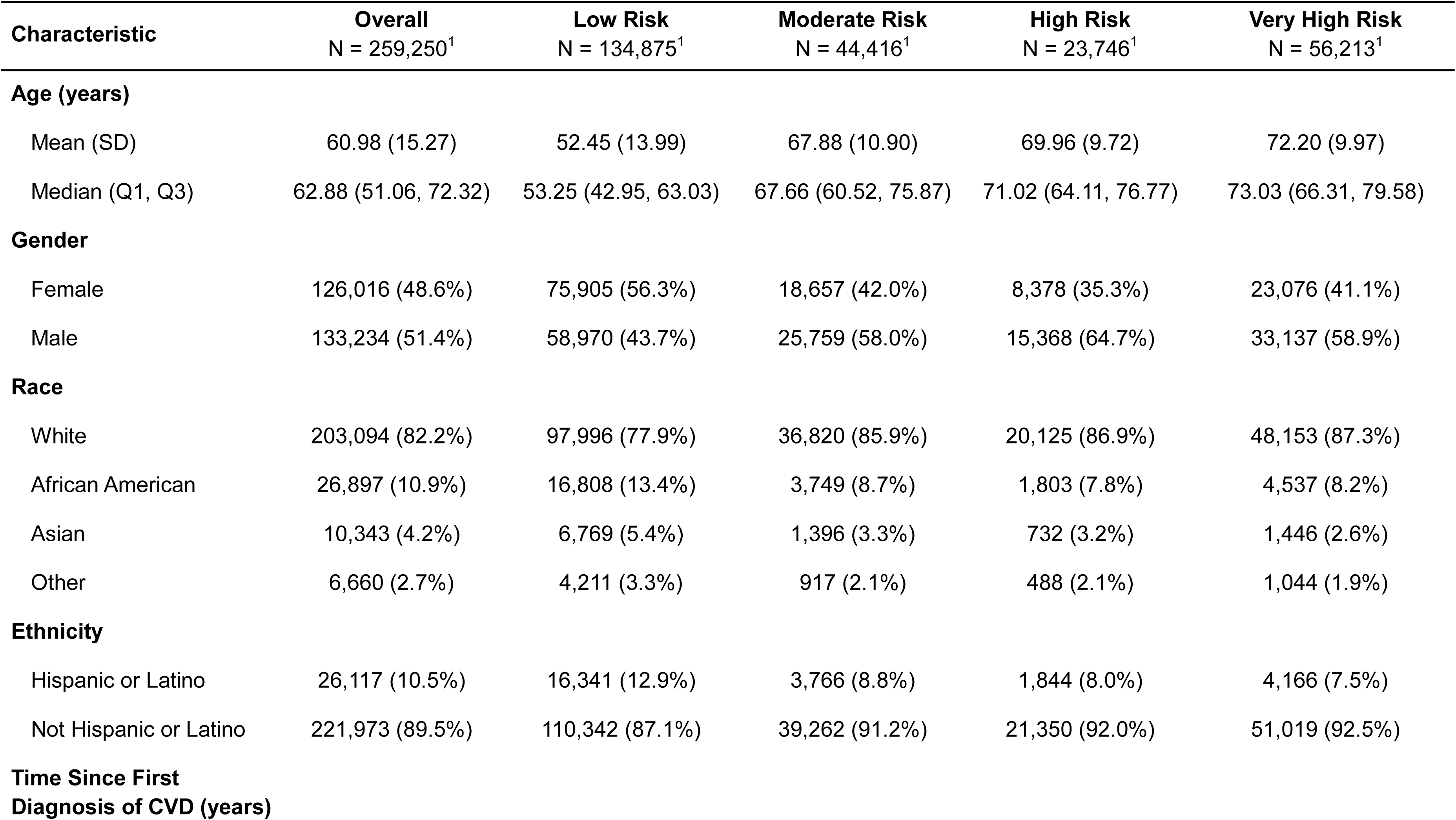
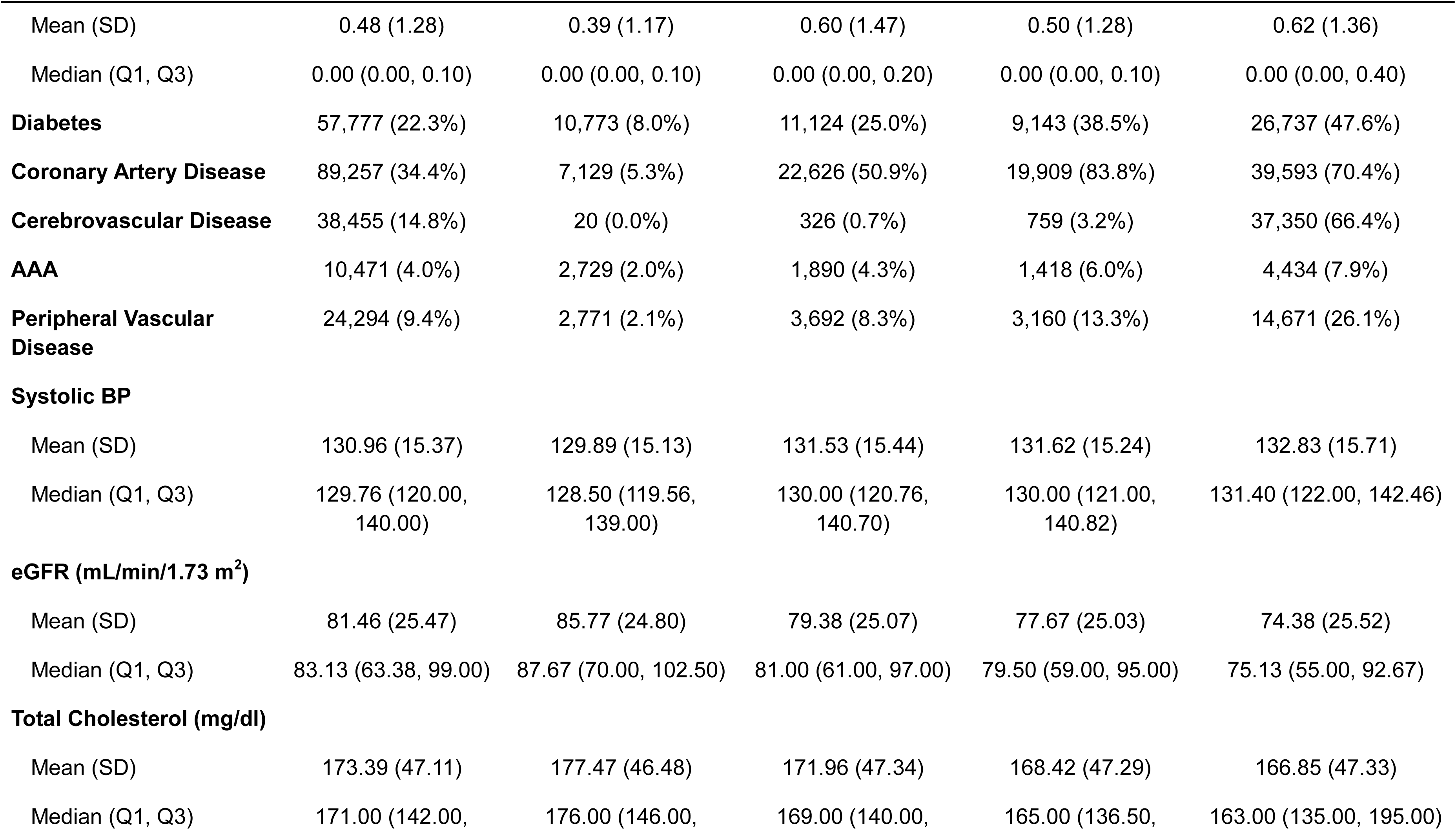
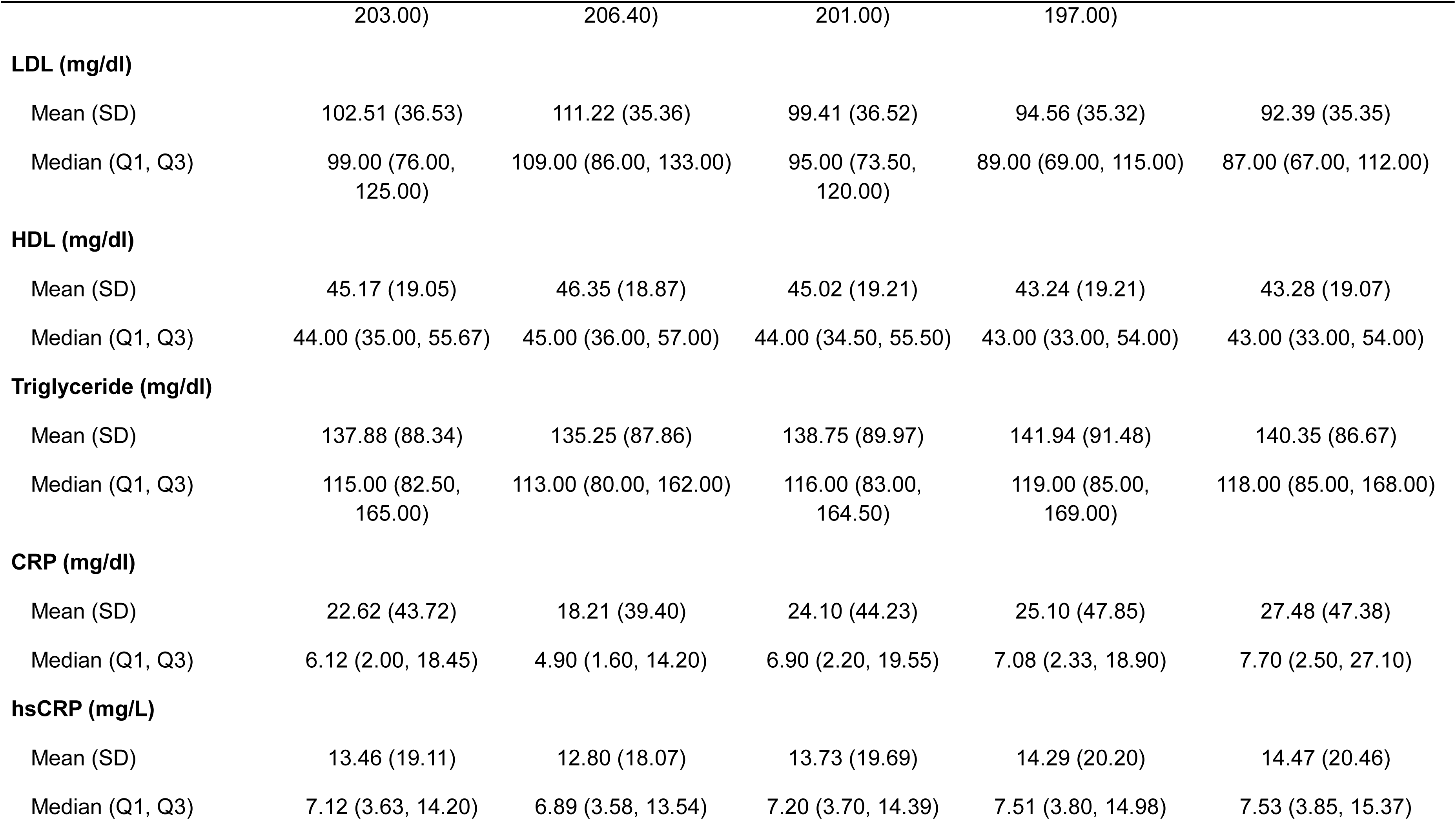
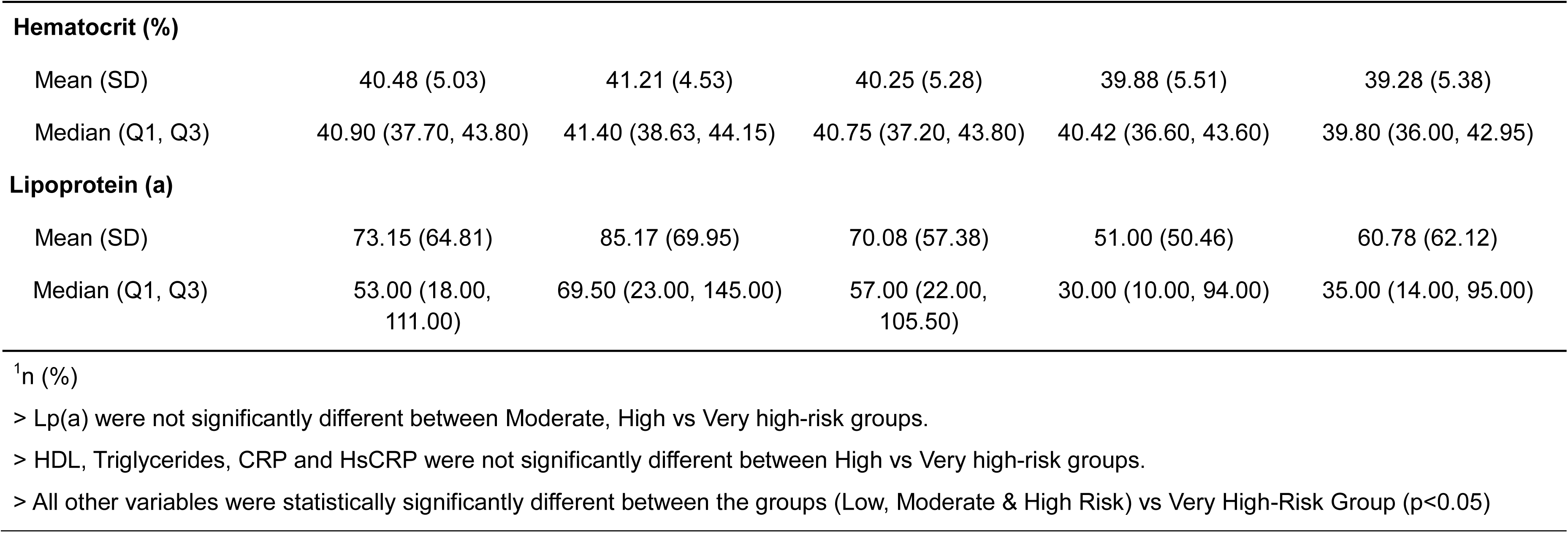
Baseline Characteristics: SMART Risk Score Variables: hsCRP – high-sensitive c-reactive protein; eGFR – estimated glomerular filtration rate; HDL – high density lipoprotein; BMI – body mass index; LDL – low density lipoprotein; SD – standard deviation.

The AFT model used to predict MACE based on SMART risk score variables is presented as **Table 2**. **Table 3(a)** summarizes model performance metrics, including measures of discrimination, concordance, and goodness-of-fit across the development, test, and overall cohorts.

**Table 2:**
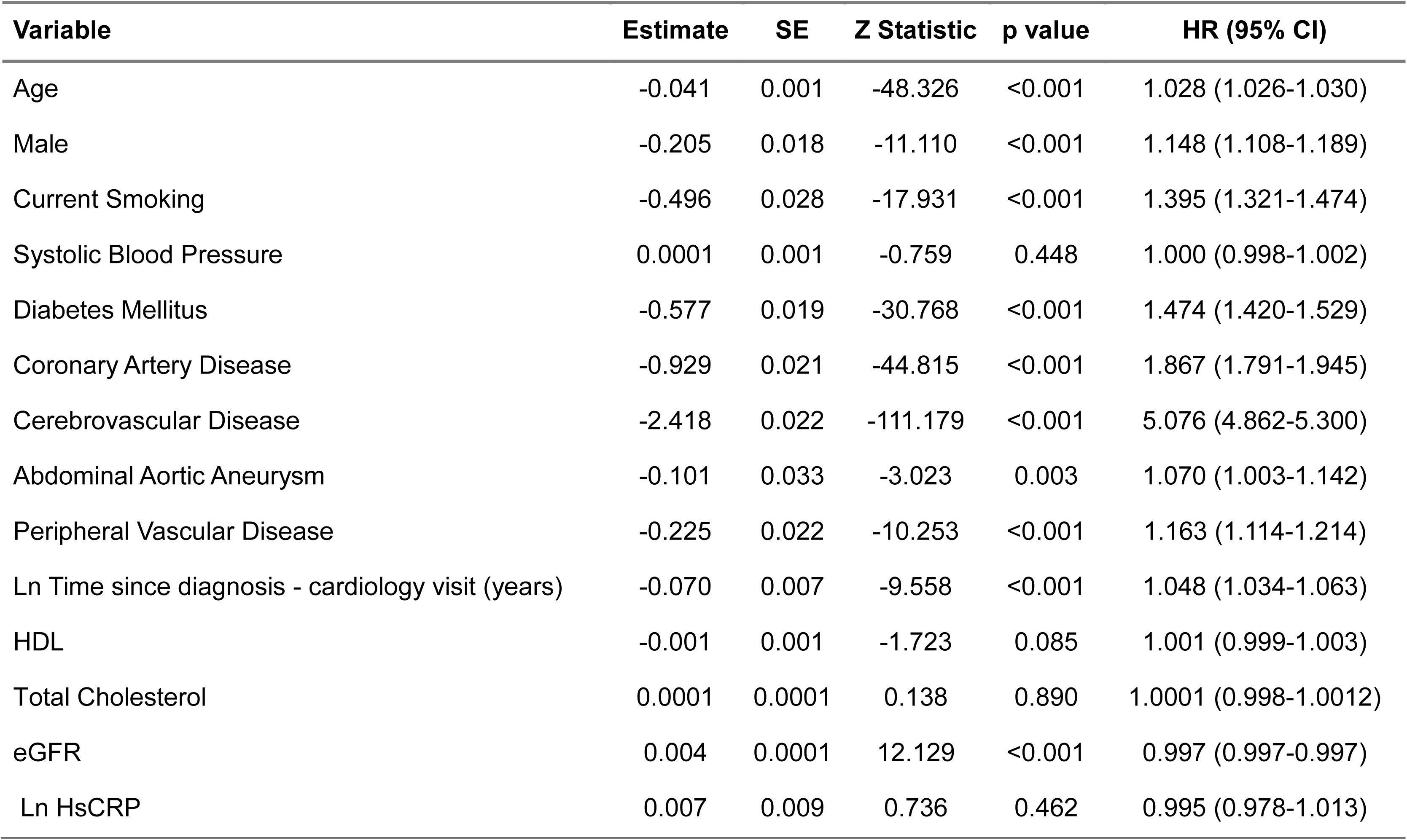
Accelerated Failure Time Model: Model Coefficients and Hazard Rates (HR) with 95% confidence intervals (CI): SE – standard error; HDL – high density lipoprotein; eGFR – estimated glomerular filtration rate; Ln: logarithmic value; hsCRP – high-sensitivity C-reactive protein. Scale=1.52; HR = exp((-Estimate)/Scale).

**Table 3:**
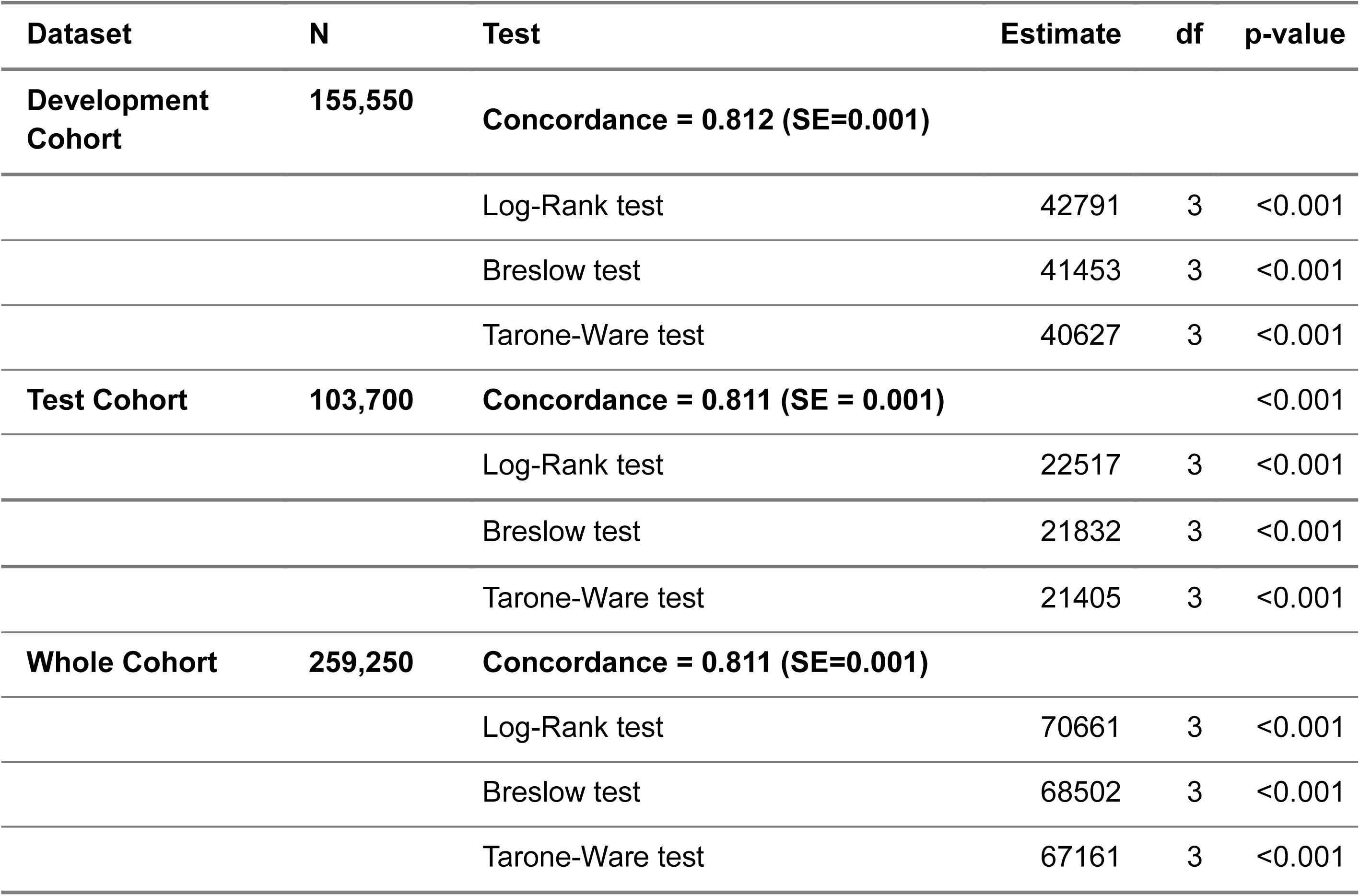
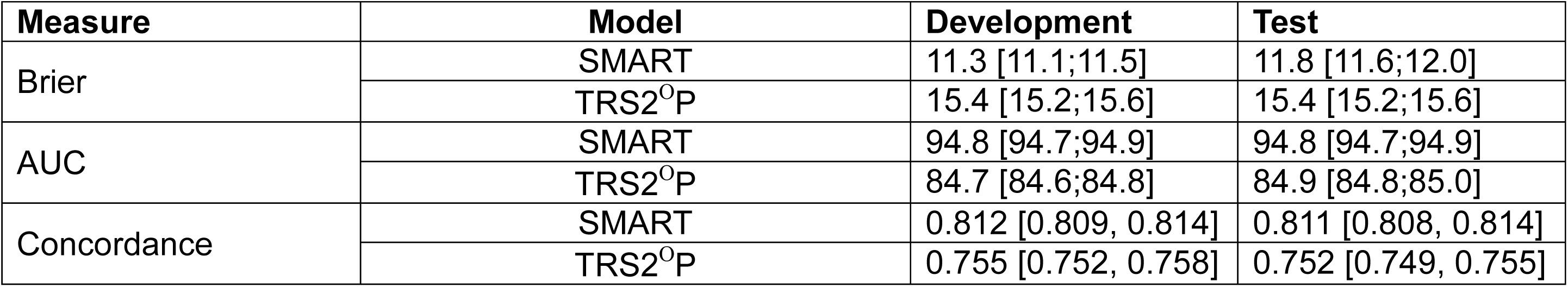
Model Performance, Concordance and Fit Tests: (a) Summary Statistics of Model Performance in the three cohorts (b) Comparison of SMART Risk score and TRS2P score.

The model demonstrated strong discriminatory ability, with a c-statistic of 0.811 in the overall cohort. Exclusion of hsCRP from the AFT model did not meaningfully alter performance metrics, including AIC, concordance, or log-likelihood values, across any dataset (development, validation, or combined), as shown in **Supplementary Table 1**. Both versions of the model, with and without hsCRP exhibited nearly identical performance across all evaluated criteria.

Comparative analysis revealed that the SMART Risk Score consistently outperformed the TRS2°P score. Specifically, the SMART score demonstrated a lower Brier score (11.3 vs. 15.4), higher area under the curve (AUC: 94.8% vs. 84.7%), and greater concordance (0.812 vs. 0.755), indicating superior predictive accuracy and overall model performance **[Table 3(c)]**.

The calibration plot of 10-year predicted versus observed event-free survival (i.e.,1-risk) for the test cohort with predicted vs observed line having minimal deviation with respect to the line of identity **(Figure 2)**. The deviance of the predicted and observed outcomes was significant in the test cohort (D’Agostino Nam test: p<0.001), attributed to a large sample size.

The event rates of MACE and its components are described across predefined risk categories, as illustrated in **Figure 3**. **Figures 4(a)** and **4(b)** present K–M survival curves for the development and test cohorts, respectively, stratified by the four SMART risk categories. These curves demonstrate that observed survival patterns are consistent with model-based risk stratification. K–M event rates for MACE and its individual components, along with mean time to MACE, are detailed in **Tables 4(a)**, **4(b)**, and **4(c)** for the overall cohort, as well as the development and test cohorts, respectively. The very high-risk group, as classified by the SMART risk score, exhibited the highest MACE rates, primarily driven by ischemic stroke and all-cause mortality, and had the shortest mean time to MACE. Comparisons between the very high-risk group and the other risk categories (low, moderate, and high) revealed statistically significant differences in all assessed variables (p<0.05).

**Figure 3:**
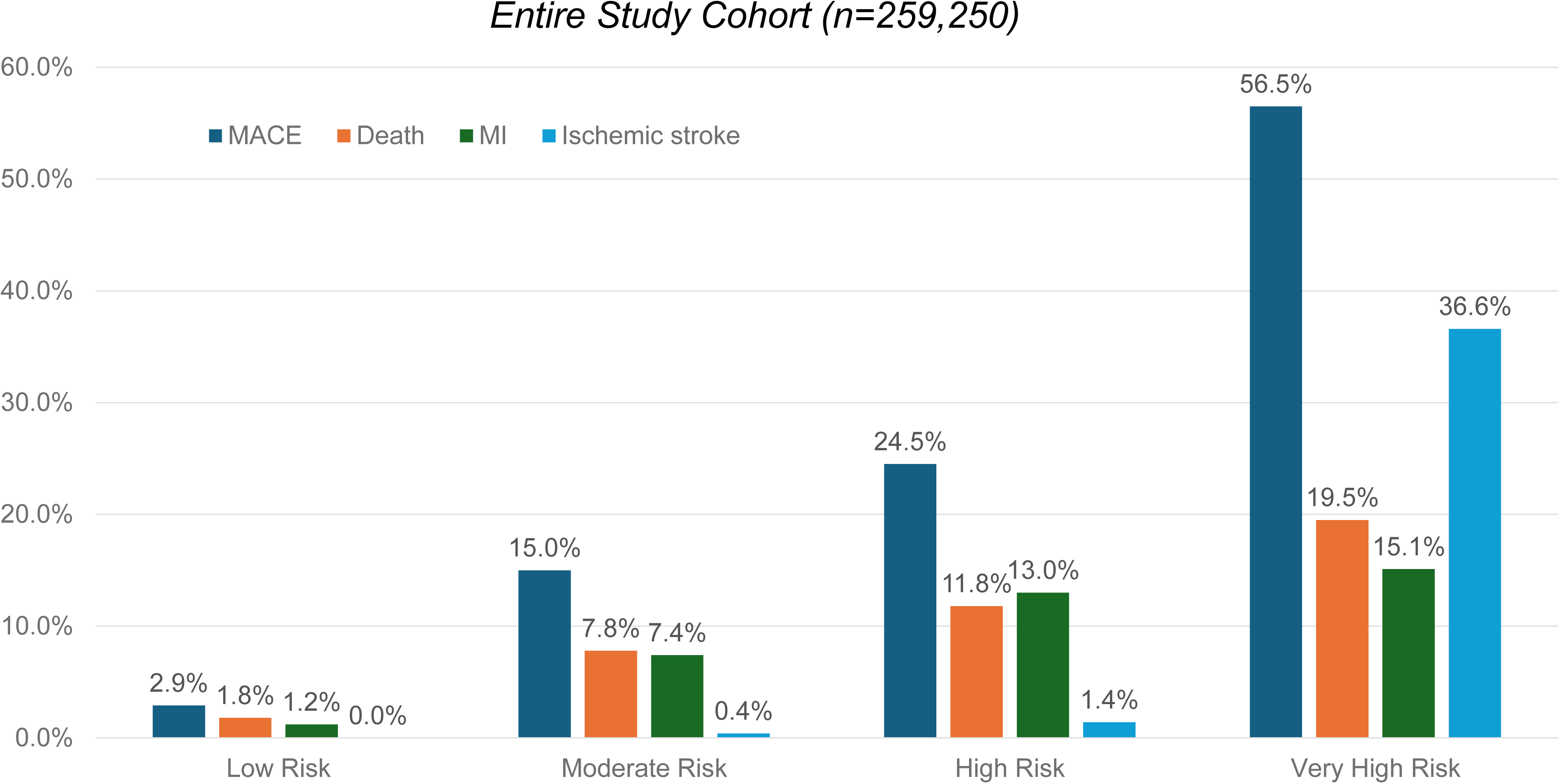
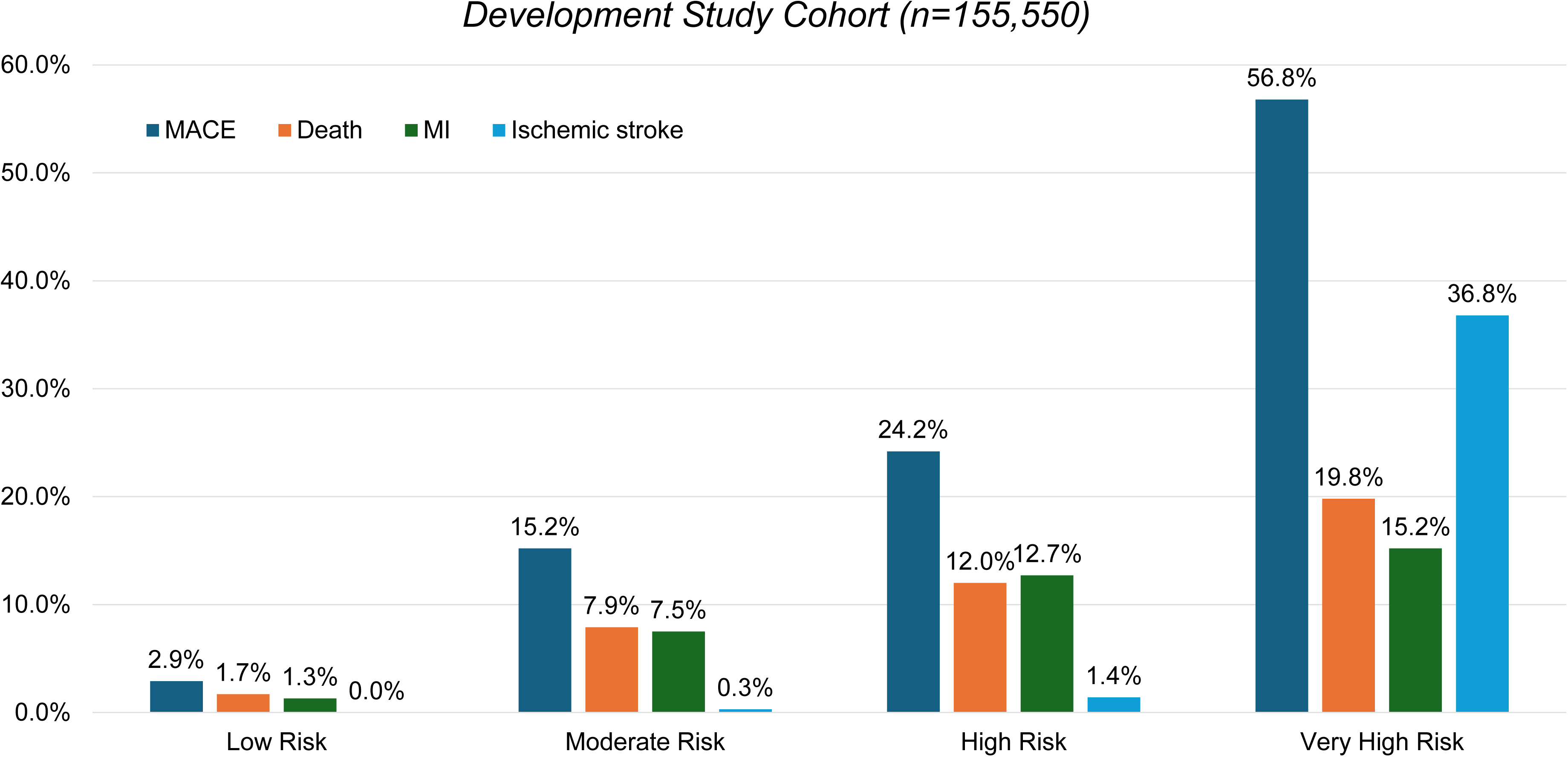
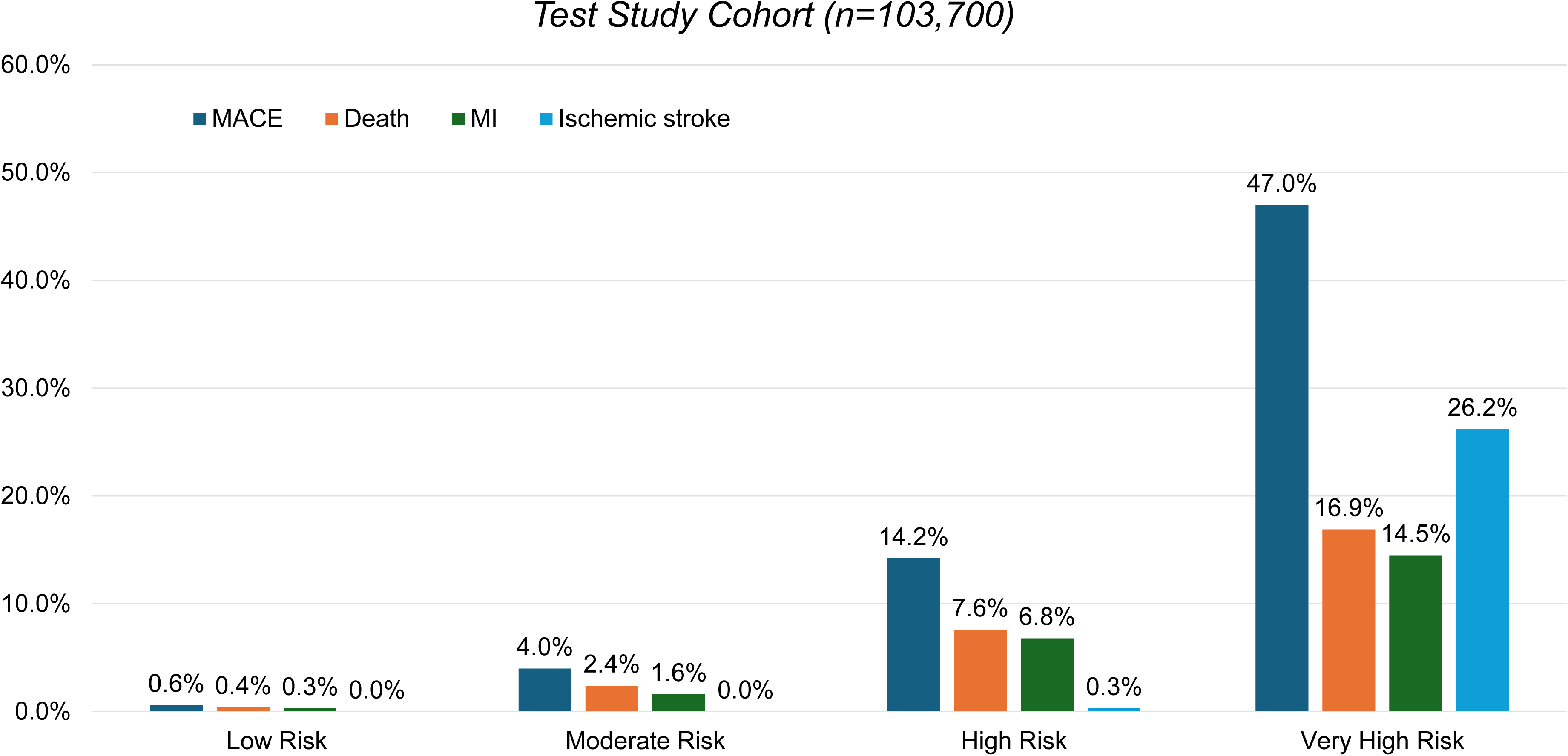
Cardiovascular Outcomes in the SMART Risk Score Groups: (a) Entire cohort; (b) Development cohort; (c) Test Cohort. MACE – major adverse cardiovascular event; MI: non-fatal myocardial infarction.

**Figure 4:**
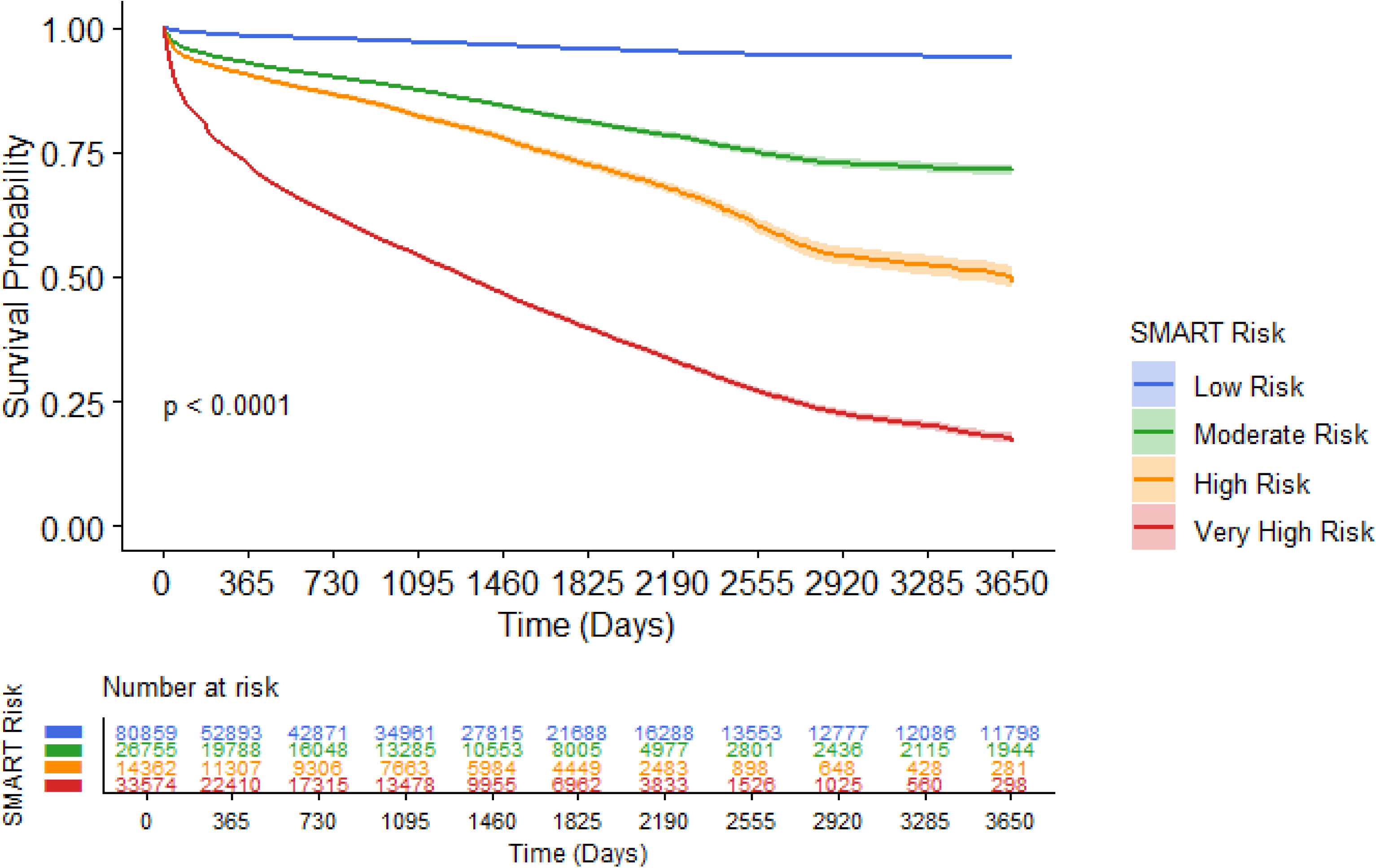
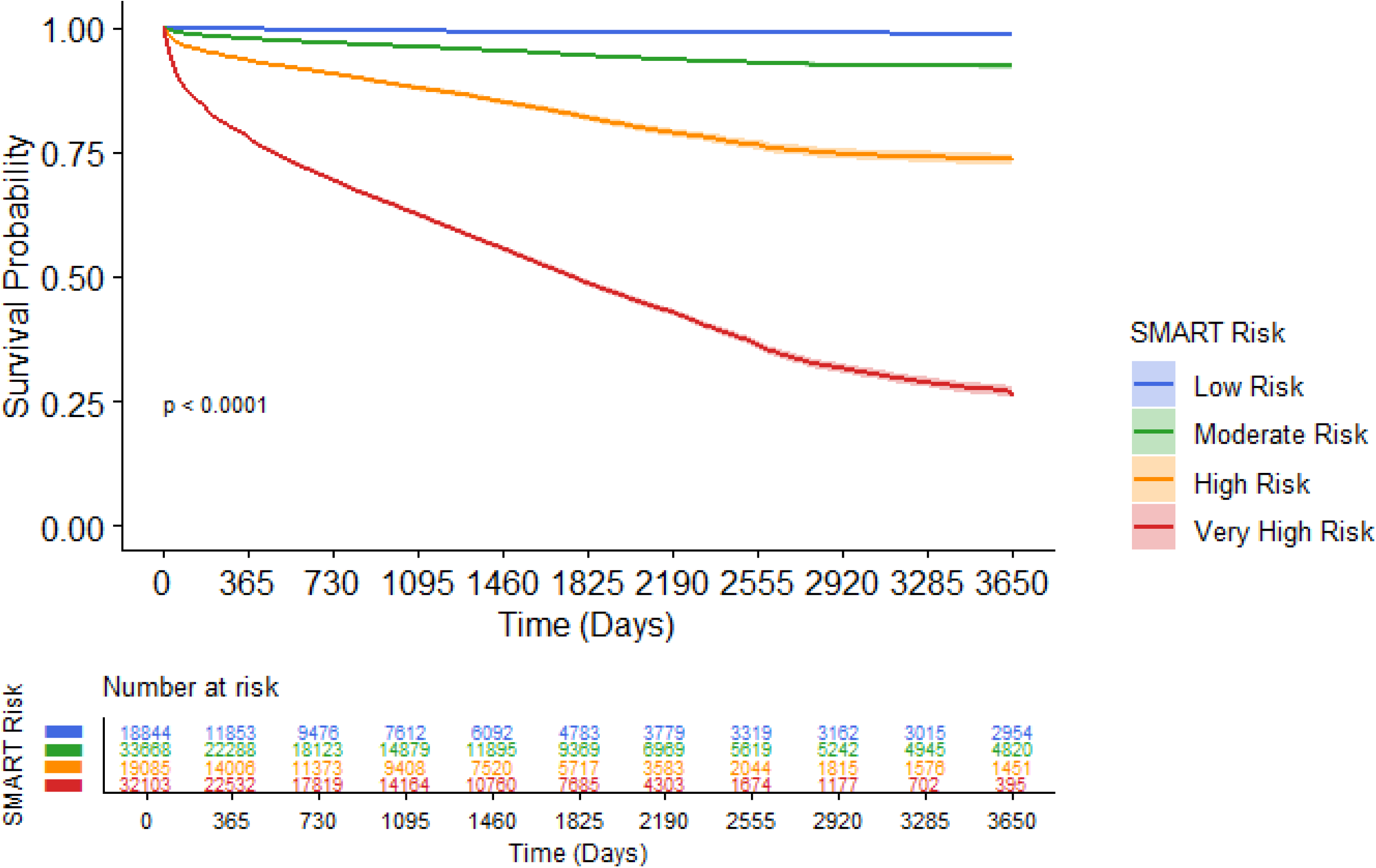
Kaplan-Meier Analysis for MACE: (a) Development cohort; (b) Test cohort. MACE – major adverse cardiovascular event.

**Table 4:**
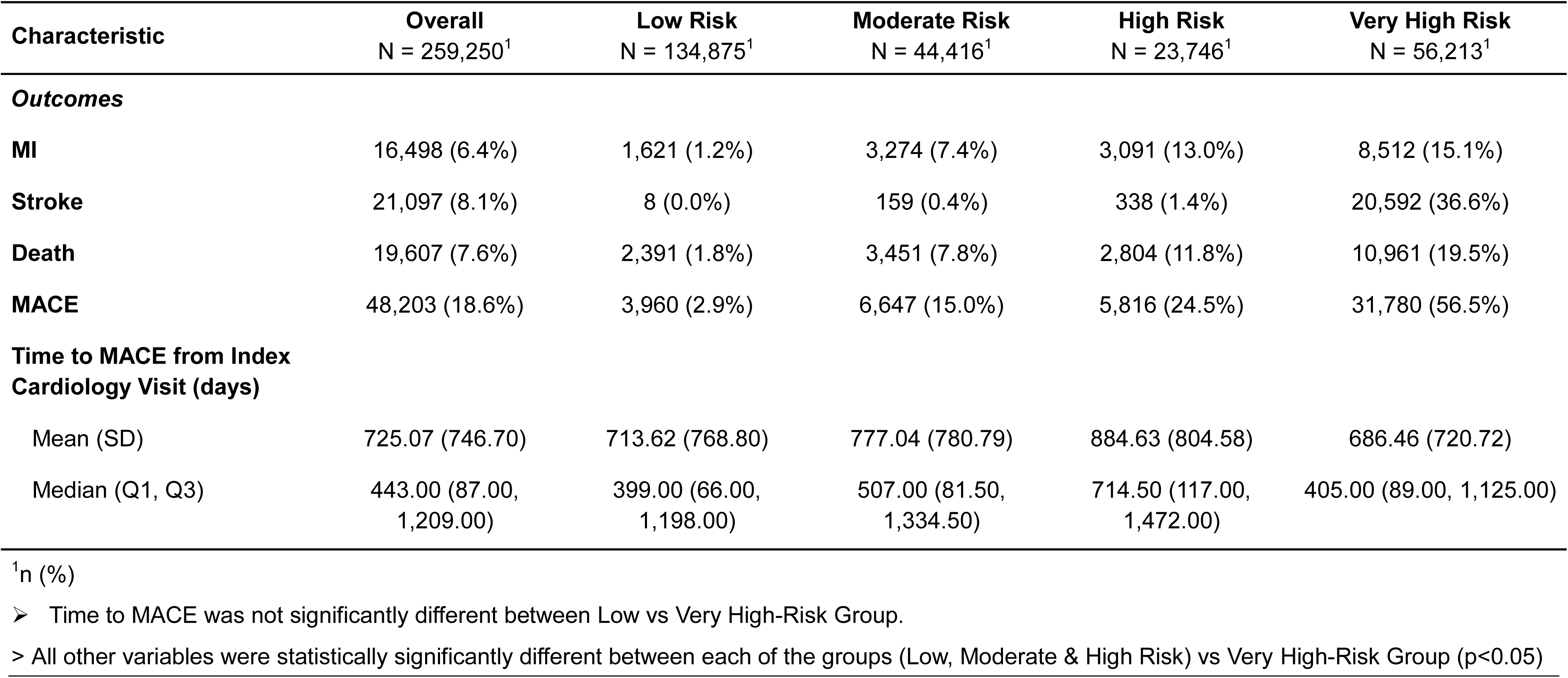
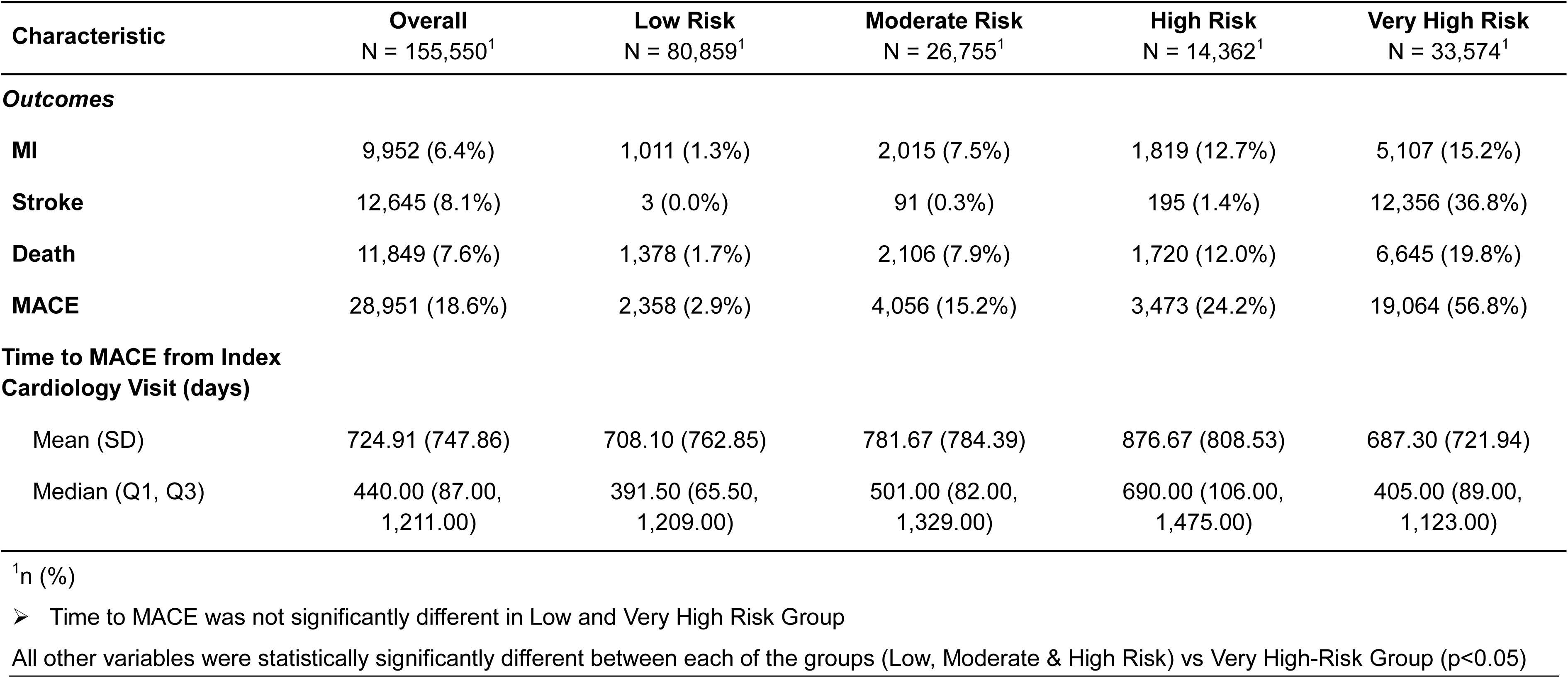
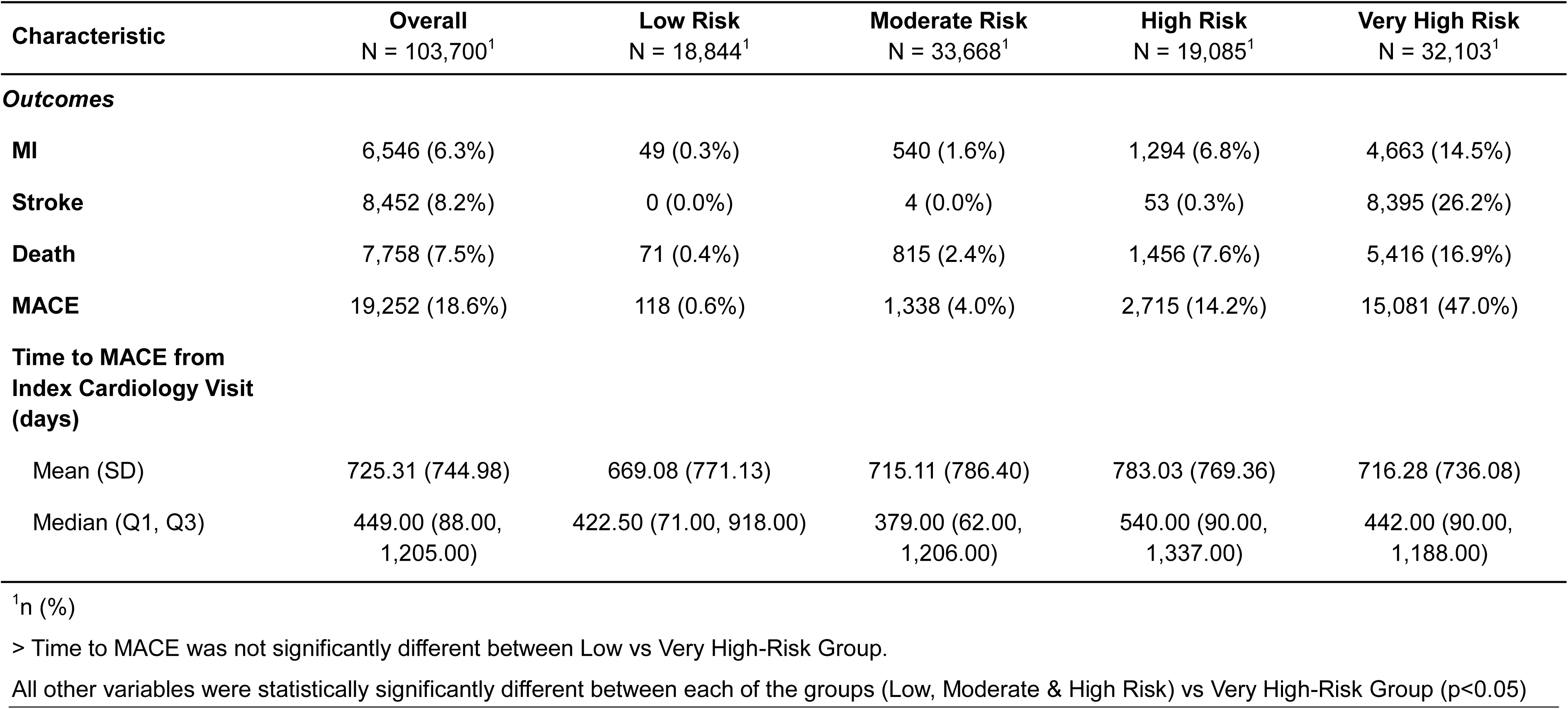
(a) Kaplan-Meir MACE Rates and Time to MACE for the Overall Cohort. (b) Kaplan-Meir MACE Rates and Time to MACE for the Development Cohort. (c) Kaplan-Meir MACE Rates and Time to MACE for the Test Cohort.

In decision curve analysis, compared with the TRS2°P score, the SMART risk score would detect an additional 110 events per 1000 patients screened at a risk threshold of 25% in the test cohort **Table 5b**.The net clinical benefit of SMART risk score (vs. TRS2°P) was even greater at higher risk thresholds of 37.5% and ≥50% as shown in **Figure 5**. A similar pattern of net clinical benefit for the SMART risk score was observed in the development cohort **Table 5a**.

**Figure 5:**
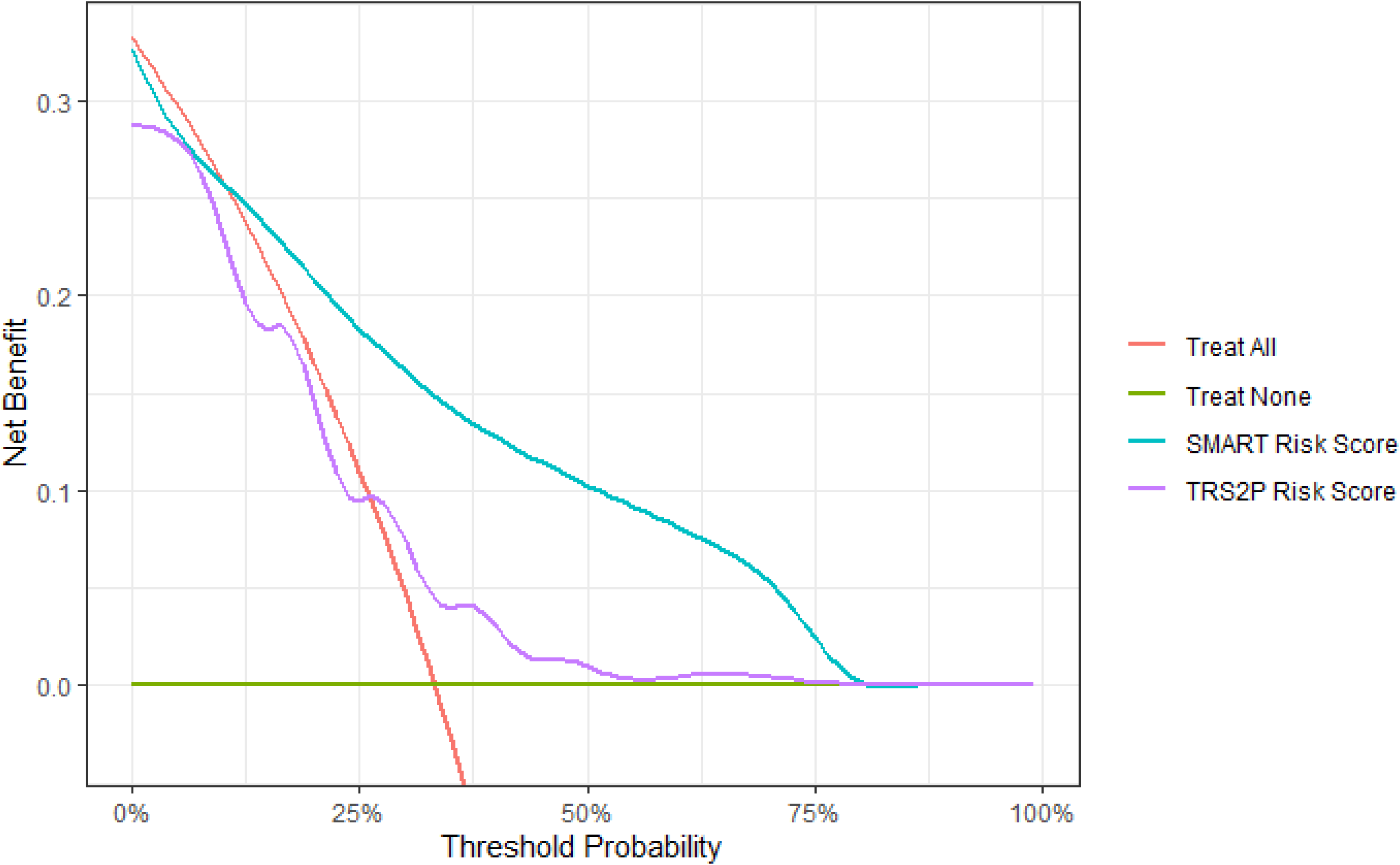
SMART Risk and TRS2°P Score Decision Analysis for Major Adverse Cardiovascular Events. For the test cohort.

**Table 5.**
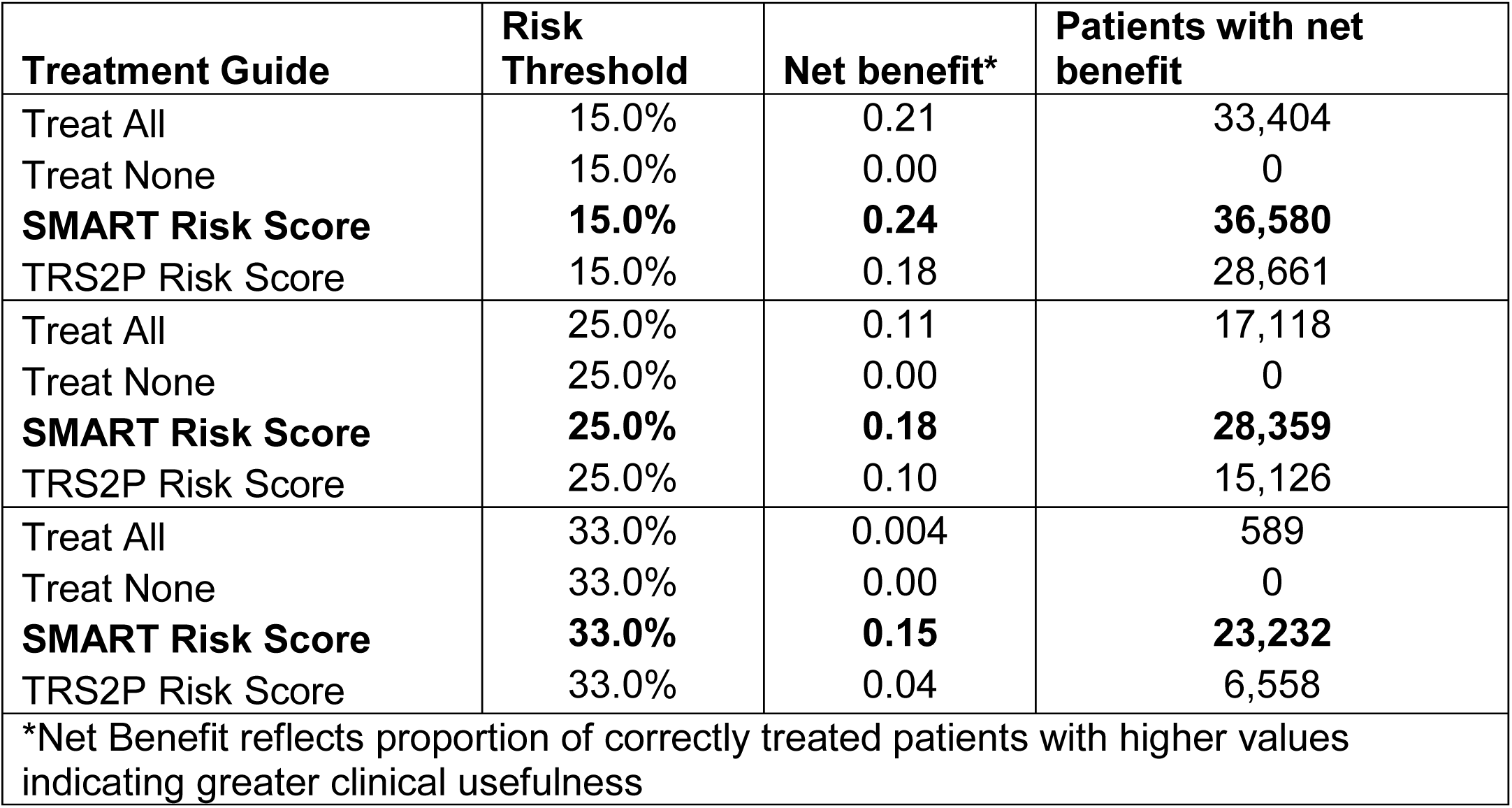
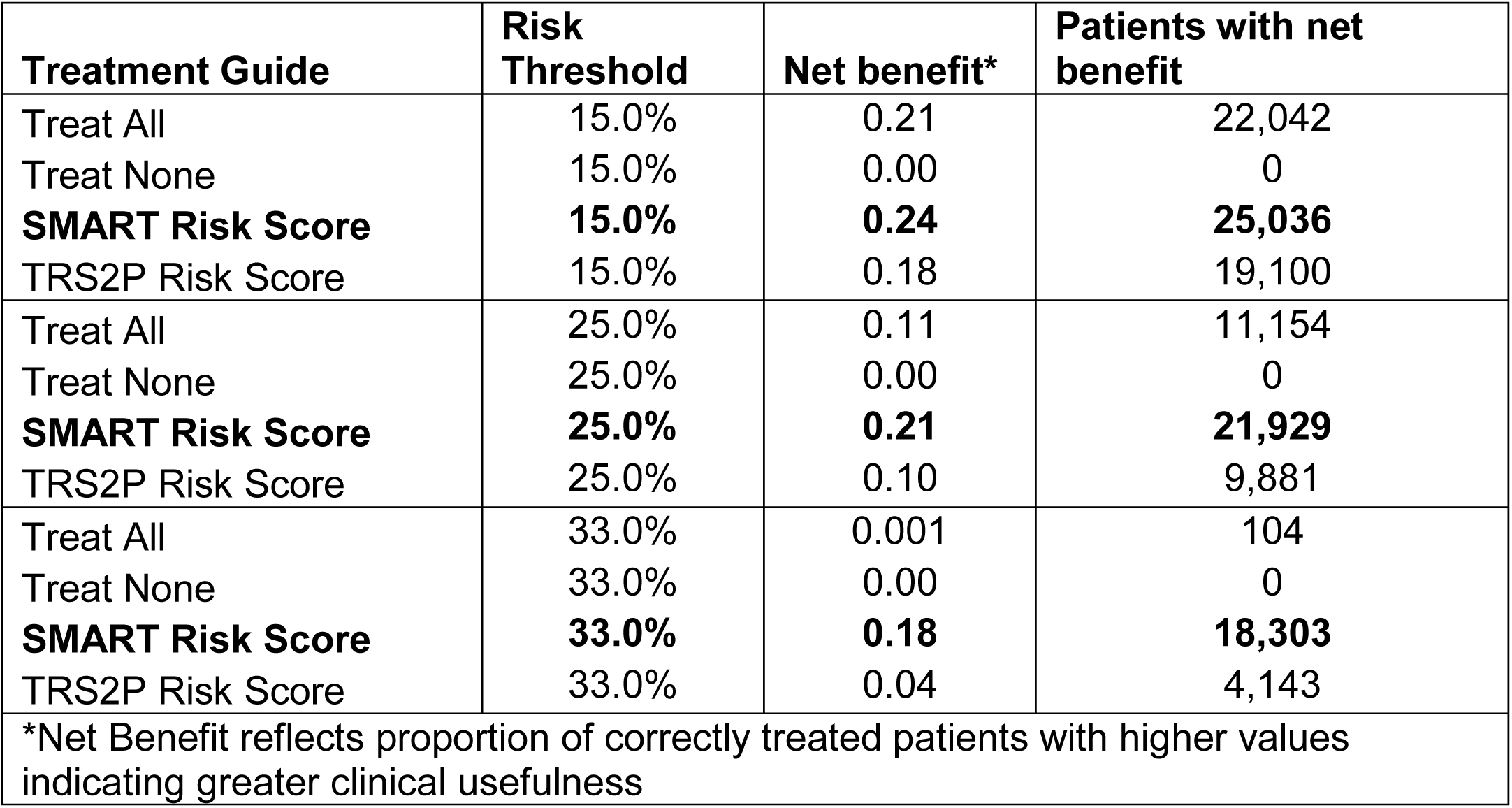
(a) Net Benefit of the SMART Risk Score for Guiding Secondary Prevention (Development Cohort) (b) Net Benefit of the SMART Risk Score for Guiding Secondary Prevention (Test Cohort):

To assess the consistency of performance across sexes, we developed separate SMART risk score models for men and women **[Supplementary Figures 1(a) and 1(b)],** race **[Supplementary Figures 2(a) and 2(b)]** and ethnicity **[Supplementary Figure 3(a) and 3(b)]** and observed consistent trends across subgroups.

## Discussion

In this large-scale study of 259,250 patients, we present robust validation of the SMART risk score as a tool for predicting long-term MACE in a real-world cohort of U.S. patients with established CVD. This is one of the largest studies to date evaluating the predictive power of the SMART risk score, which incorporates 14 clinically accessible risk predictors. By offering a comprehensive assessment across a wide spectrum of cardiovascular risk—from low to very high, the SMART risk score provides critical insights into patient stratification, facilitating the development of personalized, cost-effective prevention strategies.

Our findings validate the SMART risk score’s ability to predict MACE in both the development cohort and the test cohort. The model demonstrated a strong discriminatory capacity, with a c-statistic of 0.811 across the overall cohort, which exceeds the performance metrics observed in prior SMART risk score validations.^2, 6, 8^ Notably, our cohort included a near-equal representation of women (48.6%) and a diverse clinical sample reflective of contemporary U.S. healthcare practice.

Comparative analyses revealed that the SMART risk score outperforms other established risk models, such as the TRS2°P score, across several key metrics, including Brier score, area under the AUC, and concordance.^3^ The SMART risk score’s superior predictive accuracy, particularly in the very high-risk category, underscores its potential to better guide clinical decision-making for high-risk CVD patients.^4^

A major strength of this study is its validity within a U.S. healthcare setting, which differs significantly from the original Dutch cohort in terms of population characteristics, disease burden, and clinical management. The SMART risk score’s ability to predict MACE across different cohorts—spanning from the original Dutch cohort to the Registry of Risk factors and Event occurrence in patients with Atherothrombosis undergoing long-term Care (REACH) registry, and now to this large U.S. cohort—demonstrates its generalizability and adaptability across diverse clinical environments.^7, 8^ The findings from our study complement previous work, such as the REACH registry validation and the U.S. Veterans cohort, supporting the score’s applicability to various populations with established CVD.^5, 6, 8^

Furthermore, the decision curve analysis strengthens the clinical utility of the SMART risk score. This analysis revealed that the score offers the highest net benefit across a broad range of threshold probabilities, outperforming both the ‘treat-all’ and ‘treat-none’ strategies. ^1,2,15^ Such findings endorse the SMART risk score as a valuable tool for optimizing treatment decisions, balancing the benefit-to-harm ratio, and minimizing overtreatment—a key consideration in real-world clinical settings where precision medicine is increasingly prioritized. In this respect, the SMART risk score outperforms the TRS2°P score.

While this study highlights the SMART risk score’s potential for clinical practice, several limitations warrant consideration. Firstly, the retrospective nature of our data, derived from electronic medical records, introduces the possibility of data inaccuracies, particularly in capturing certain risk factors and medication adherence. Furthermore, although our cohort includes a substantial proportion of women, the study population is predominantly white, limiting the generalizability of the findings to more racially and ethnically diverse groups. Ongoing and future efforts to validate the SMART risk score in diverse populations are essential to confirm its applicability across different demographic subgroups and healthcare settings. The c-statistic of 0.811 achieved in this study marks an improvement over previous SMART risk score validations, which generally reported c-statistics around 0.67.^2^ While this represents a meaningful advancement, further refinement and validation of the model, particularly in external datasets, will be critical to addressing remaining uncertainties, such as the potential for calibration in populations with differing clinical profiles.

In conclusion, the SMART risk score provides an effective, reproducible method for predicting long-term cardiovascular risk in patients with established polyvascular disease. Our findings emphasize its clinical utility for improving risk stratification, guiding individualized treatment decisions, and optimizing secondary prevention strategies. Further validation in more diverse cohorts is necessary to fully understand its potential for broad clinical implementation. With additional studies, the SMART risk score has the potential to become a cornerstone of personalized cardiovascular care, driving more informed, precision-based interventions that improve long-term outcomes for patients with cardiovascular disease.

## Data Availability

We will not be able to provide access to source data.

---

**Supplementary Figure 1:**
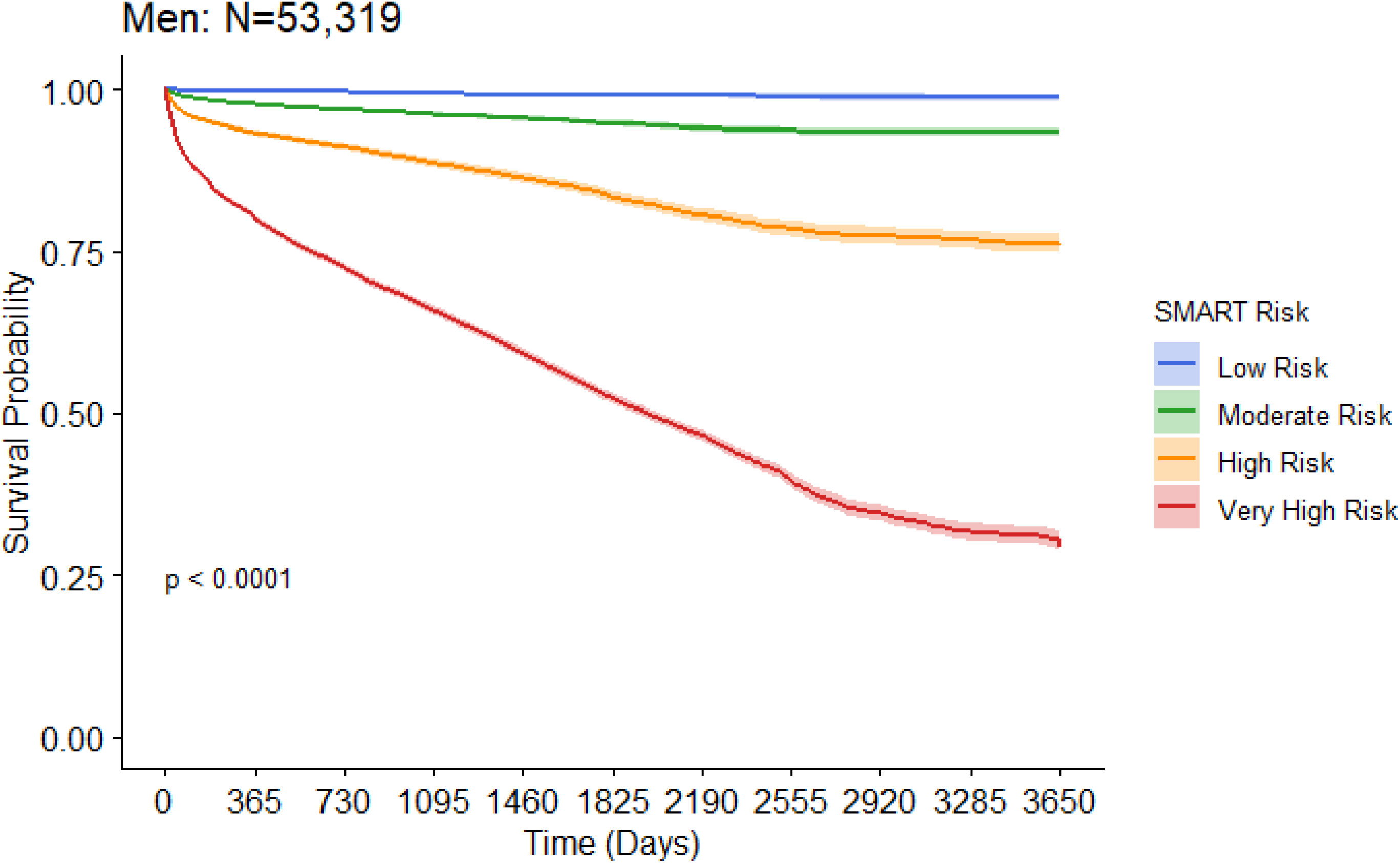
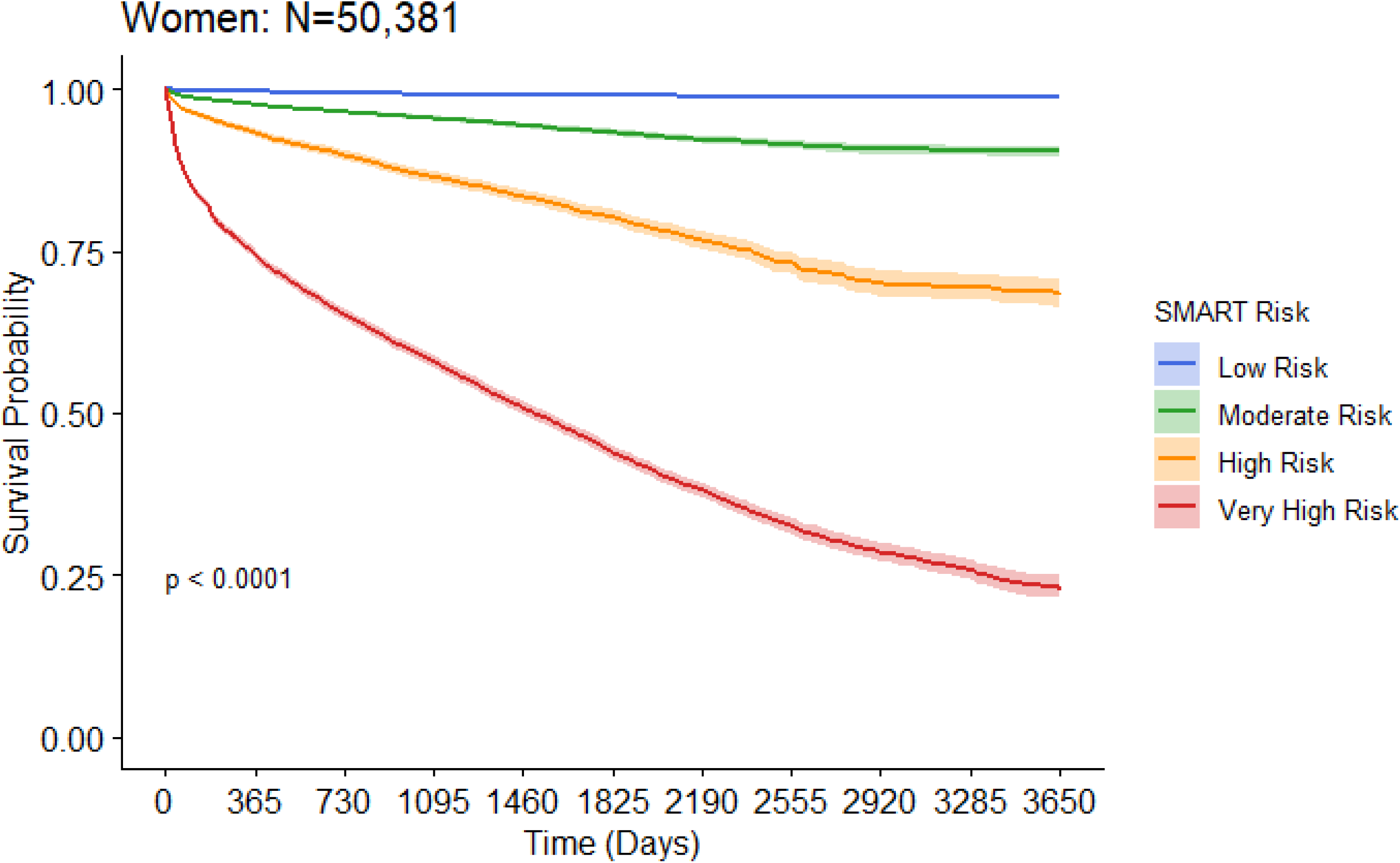
Kaplan-Meier Analysis for MACE by Gender: (a) Men; (b) Women. MACE – major adverse cardiovascular event.

**Supplementary Figure 2:**
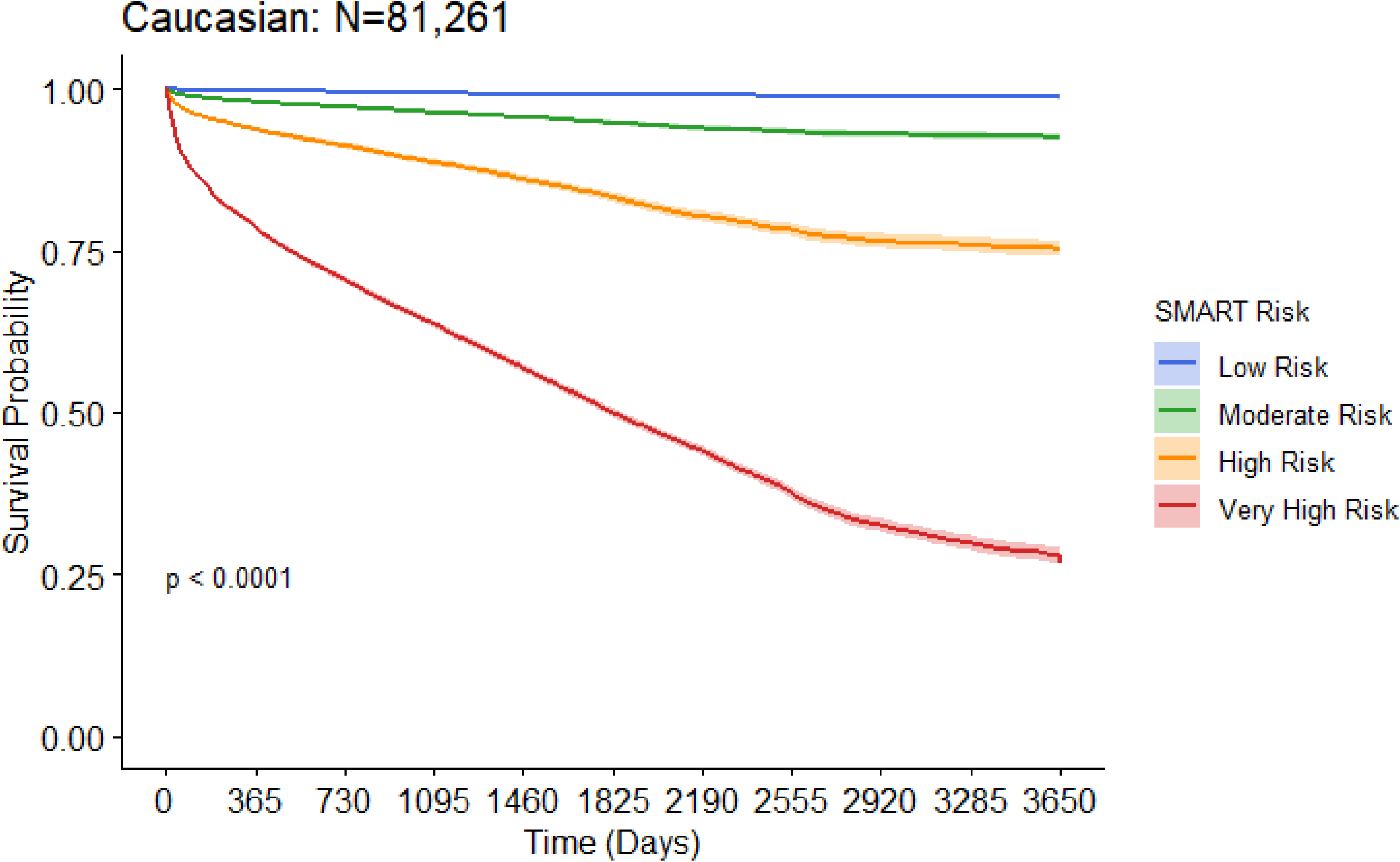
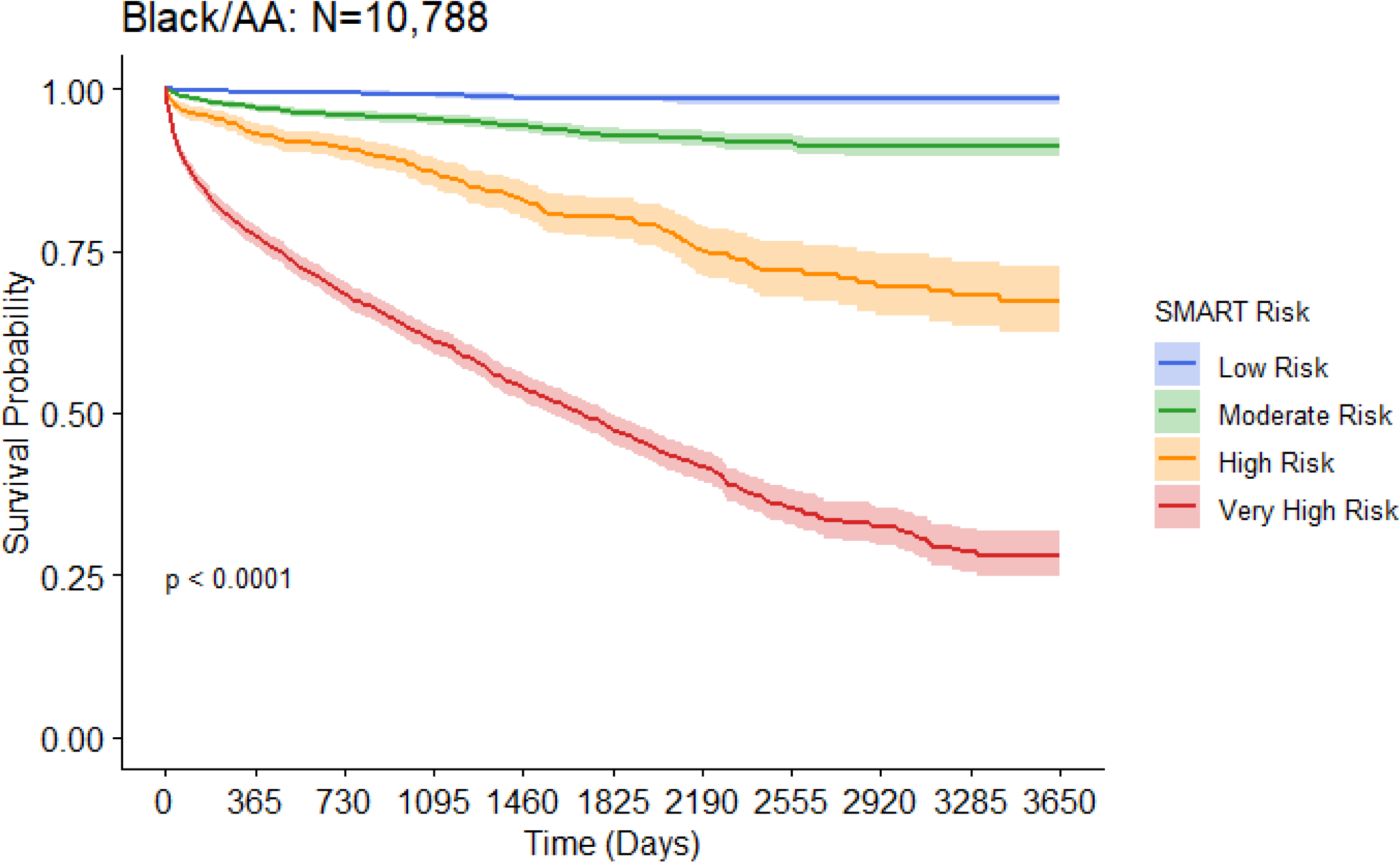
Kaplan-Meier Analysis for MACE by Race: (a) Caucasian; (b) Black/AA; MACE – major adverse cardiovascular event; AA – African American.

**Supplementary Figure 3:**
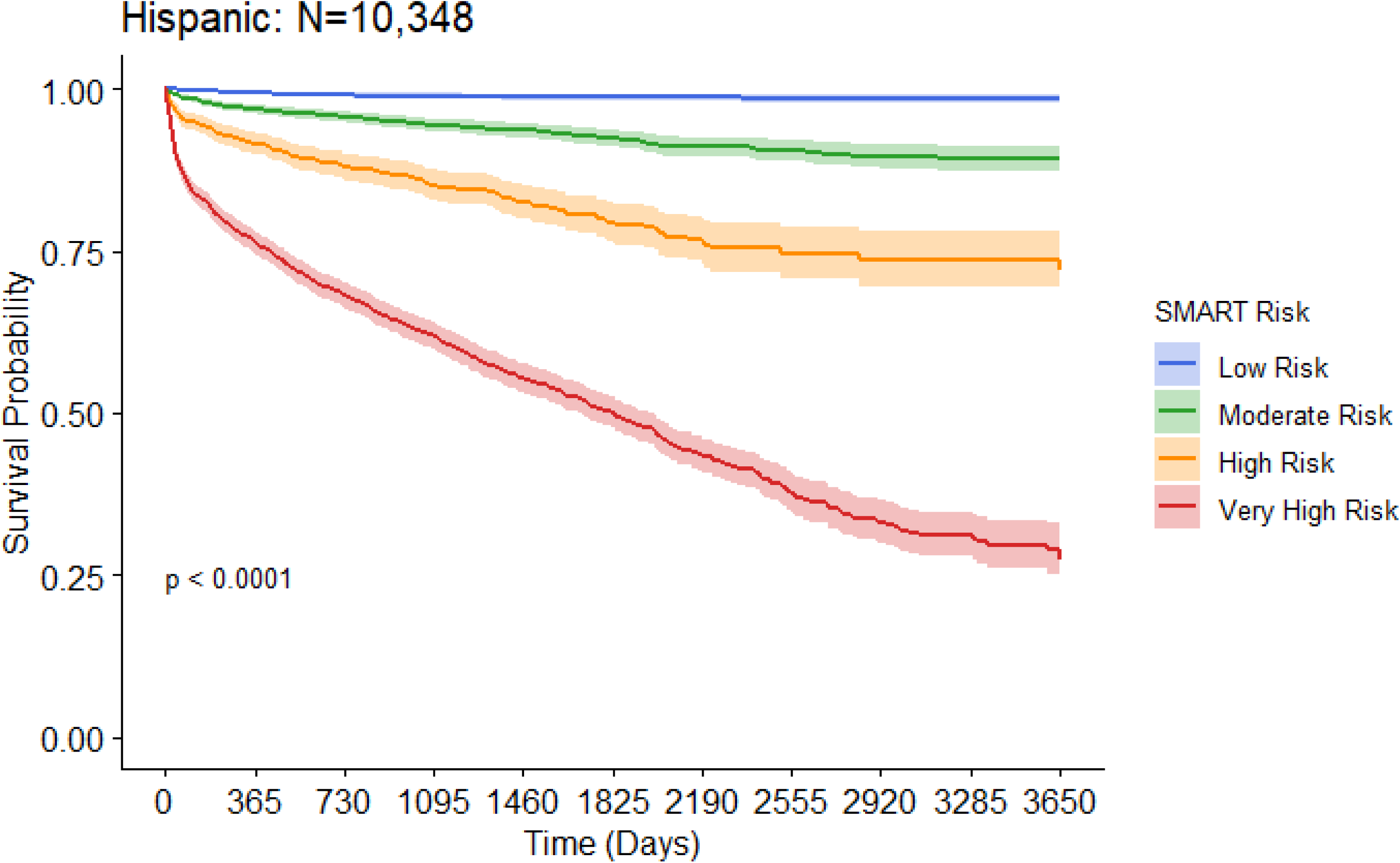
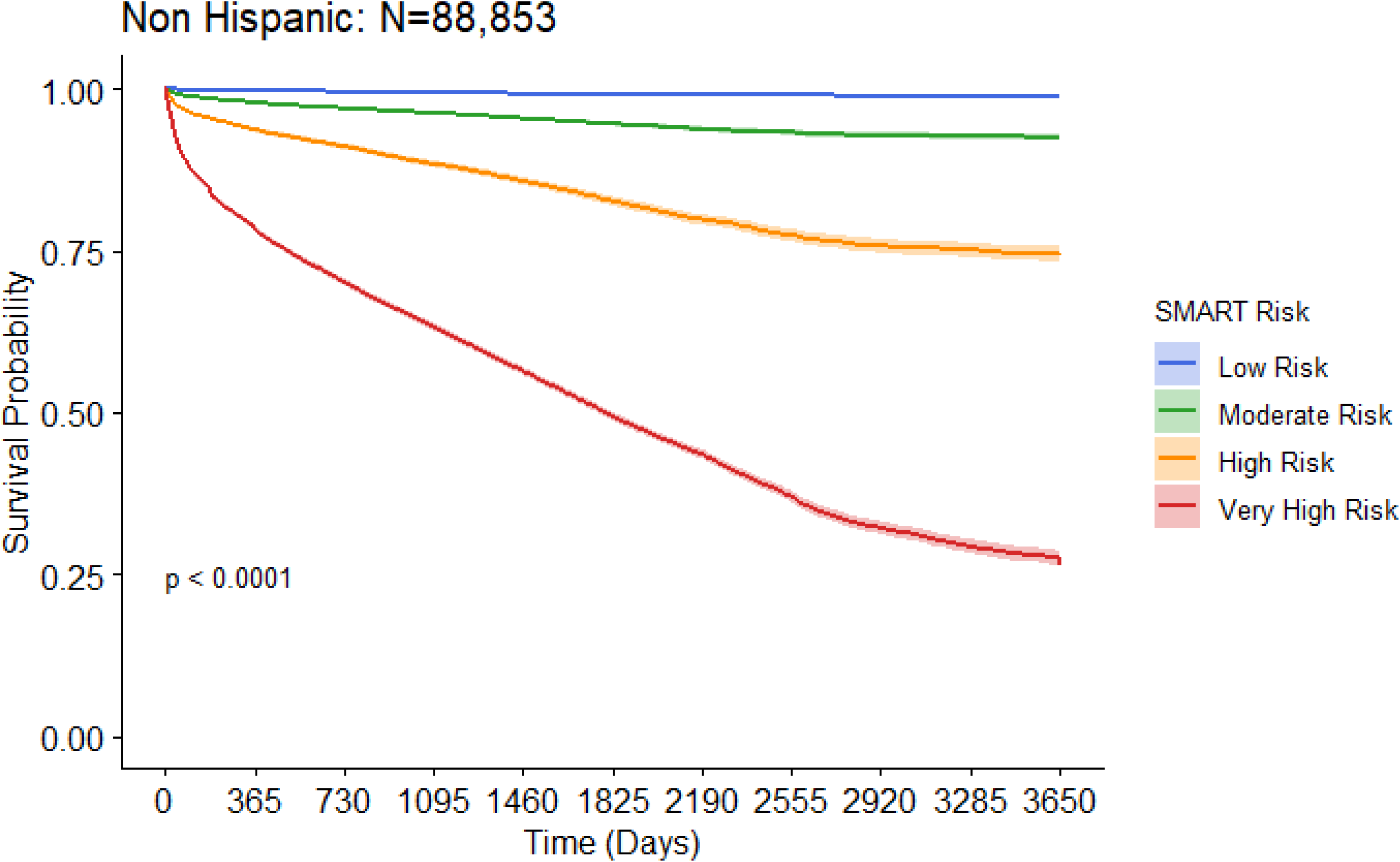
Kaplan-Meier Analysis for MACE by Ethnicity: (a) Hispanic; (b) Non-Hispanic. MACE – major adverse cardiovascular event.

**Appendix 1:** Study International Classification of Diseases (ICD) 9 and 10 and Current Procedure Terminology (CPT) codes.

**Appendix 2:** TRIPOD Guideline Adherence Document

**Central Figure:**
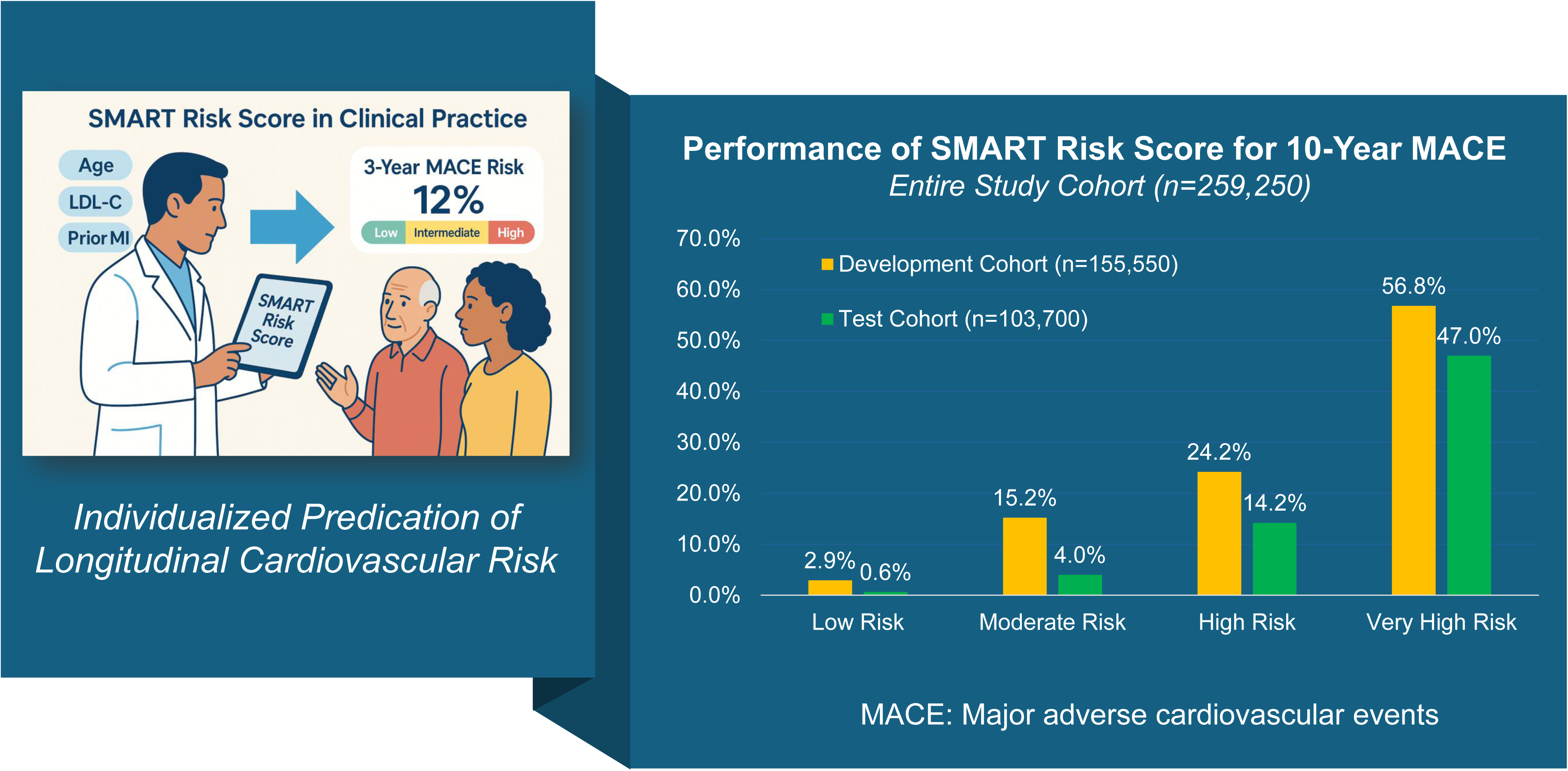
Estimating the Risk of Cardiovascular Events in U.S. Veterans Using the SMART Risk Score.

**Supplementary Table 1:** Comparison of SMART risk score model performance with or without HsCRP.

**Supplementary Table 2:** Model performance and other statistics for the Subgroups in the Test Cohort.

## Acknowledgements

We would like to acknowledge research grant support from Esperion Inc., philanthropic support from Bob and Brigitta Smith to the Baylor Heart and Vascular Lipid Clinic, and philanthropic support from Ashley and Greg Arnold to the Baylor Research Institute.

